# Genome-Wide Assessment of Pleiotropy Across >1000 Traits from Global Biobanks

**DOI:** 10.1101/2025.04.18.25326074

**Authors:** Michael G Levin, Satoshi Koyama, Jakob Woerner, David Y Zhang, Alexis Rodriguez, Tarak Nandi, Buu Truong, Sarah A Abramowitz, Hritvik Gupta, Himani Kamineni, Whitney Hornsby, Zilinghan Li, Taylor Cohron, Jennifer E Huffman, Patrick Ellinor, Dokyoon Kim, Katherine P Liao, Ravi K Madduri, Benjamin F Voight, Anurag Verma, Scott M Damrauer, Pradeep Natarajan

## Abstract

Large-scale genetic association studies have identified thousands of trait-associated risk loci, establishing the polygenic basis for common complex traits and diseases. Although prior studies suggest that many trait-associated loci are pleiotropic, the extent to which this pleiotropy reflects shared causal variants or confounding by linkage disequilibrium remains poorly characterized. To define a set of candidate loci with potentially pleiotropic associations, we performed genome-wide association study (GWAS) meta-analyses of up to 1,167 clinically relevant traits and diseases across 1,789,365 diverse individuals genetically similar to Admixed American (AMR, N_Max_ = 60,756), African (AFR, N_Max_ = 128,361), East Asian (EAS, N_Max_ = 307,465), European (EUR, N_Max_ = 1,283,907), and South Asian (SAS, N_Max_ = 8,876) reference populations from the VA Million Veteran Program (MVP), UK Biobank (UKB), FinnGen, Biobank Japan (BBJ), Tohoku Medical Megabank (ToMMo), and Korean Genome and Epidemiology Study (KoGES). We identified 27,193 genome-wide significant locus-trait pairs (1MB region with P_GWAMA_ < 5 × 10^−8^) in within-population analysis and 29,139 in multi-population analysis (P_MR-MEGA_ < 5 × 10^−8^). Among these, 11.5% (n = 3,149) of locus-trait pairs in population-wise and 6.4% (n = 1,875) in multi-population analyses did not reach genome-wide significance in previously published GWAS. In aggregate, the genome-wide significant loci fell within 2,624 non-overlapping autosomal genomic windows on average ∼600kb in size. Each locus contained genome-wide significant signals for a median of 6 traits (IQR 2 to 18), including 2,110 (80%) pleiotropic loci associated with >1 trait. Multi-trait colocalization identified 1,902 (72%) loci with high-confidence (posterior probability > 0.9) evidence of a shared causal variant across two or more traits. Variants in pleiotropic loci were significantly enriched for a broad spectrum of functional annotations compared to non-pleiotropic counterparts. Polygenic scores (PGS) developed from these data generally improved prediction compared to existing PGS, and were broadly associated with both primary and pleiotropic phenotypes. These results provide a contemporary map of genetic pleiotropy across the spectrum of human traits/diseases and diverse genetic backgrounds.

## INTRODUCTION

Over the past 25 years, genome-wide association studies (GWAS) have revealed the complex genetic architecture of common human traits and diseases. These studies have uncovered new mechanisms of disease,^1^ identified therapeutic targets,^2^ and enabled genomic approaches to risk prediction^3^. Interrogations of published GWAS summary statistics have found that the majority of genetic variation identified by GWAS influences more than one trait, indicating that pleiotropic genetic signals are potentially pervasive.^4^ However, limitations of prior analyses of pleiotropy include (1) absence or limited inclusion of non-European populations and (2) limited adequately powered GWAS across the phenome.^5^ Recently, biobanks linking genomic data with electronic health records among large, diverse populations have provided new insights into the genetic architecture of a larger breadth of human traits and diseases across a broader spectrum of genetic backgrounds.^6,7^

To comprehensively evaluate the extent of pleiotropic signals across populations, we systematically performed GWAS meta-analysis for up to 1,167 traits/diseases across populations similar to Admixed American (AMR, N_Max_ = 59,894), African (AFR, N_Max_ = 128,206), East Asian (EAS, N_Max_ = 293,866), and European (EUR, N_Max_ = 1,282,152) reference populations across 1,789,365 participants of the VA Million Veteran Program (MVP),^6,8^ UK Biobank (UKB),^9^ FinnGen,^10^ Biobank Japan (BBJ),^11^ Tohoku Medical Megabank (ToMMo),^12^ and Korean Genome and Epidemiology Study (KoGES),^13^ with replication in 206,383 participants of the All Of Us Research Program (AoU).^7^ The results of these analyses were then used to 1) identify trait-associated loci that did not reach the genome wide significance threshold in individual studies, 2) characterize pleiotropy at the variant, locus, and trait levels across populations; and 3) perform phenome-wide genetic prediction.

## RESULTS

### GWAS meta-analysis, fine-mapping, and replication

We combined phenome-wide GWAS results from the VA Million Veteran Program (MVP), UK Biobank (UKB), FinnGen, and Biobank Japan (BBJ).^6,9–11^ We also curated quantitative trait GWAS results, generally comprising anthropometrics, vital signs, and laboratory values, from these cohorts and additional EAS cohorts [Tohoku Medical Megabank (ToMMo) and Korean genome and epidemiology study (KoGES), **Figure 1a**].^12,13^ We performed fixed-effect meta-analysis for within-population studies of 1,167 outcomes (**Supplementary Table 1,2,3**) and meta-regression using MR-MEGA for multi-population studies of 558 outcomes (**Supplementary Table 4**). We observed well-controlled test statistics in within-population analysis [median Lambda GC of 1.048 (Interquartile Range; IQR 1.021 - 1.108), median attenuation ratio of 6.47% (IQR 1.08% - 13.7%) by LD score regression (**Extended Data Figure 1**)] and multi-population analysis [median Lambda GC of 1.087 (IQR 1.043 - 1.182)]. In within-population meta-analyses, across 852 traits, we identified 27,193 genome-wide significant (GWS) locus-trait pairs defined as a region at least 1MB in length including at least one variant with *P*_GWAMA_ < 5 × 10^−8^ (**Supplementary Figure 1a, Supplementary Table 5**). In multi-population analysis, we identified 25,593 GWS locus-trait pairs with *P*_MR-MEGA_ < 5 × 10^−8^, across 472 traits (**Figure 1b, 1c, and 1d, Supplementary Table 6**). To prioritize putative causal variants, we conducted genetic fine-mapping based on the Bayes factor (**Supplementary Methods**). We identified 19,325 locus-trait pairs with a 95% credible set containing ≤ 5 variants, among which 7,187 loci had a single variant in the credible set (PIP > 0.95). The average 95% credible set size was smallest in the AFR meta-analysis (**Extended Data Figure 2b and 2c**), likely related to known relatively smaller linkage disequilibrium block sizes.^14^

**Figure 1.**
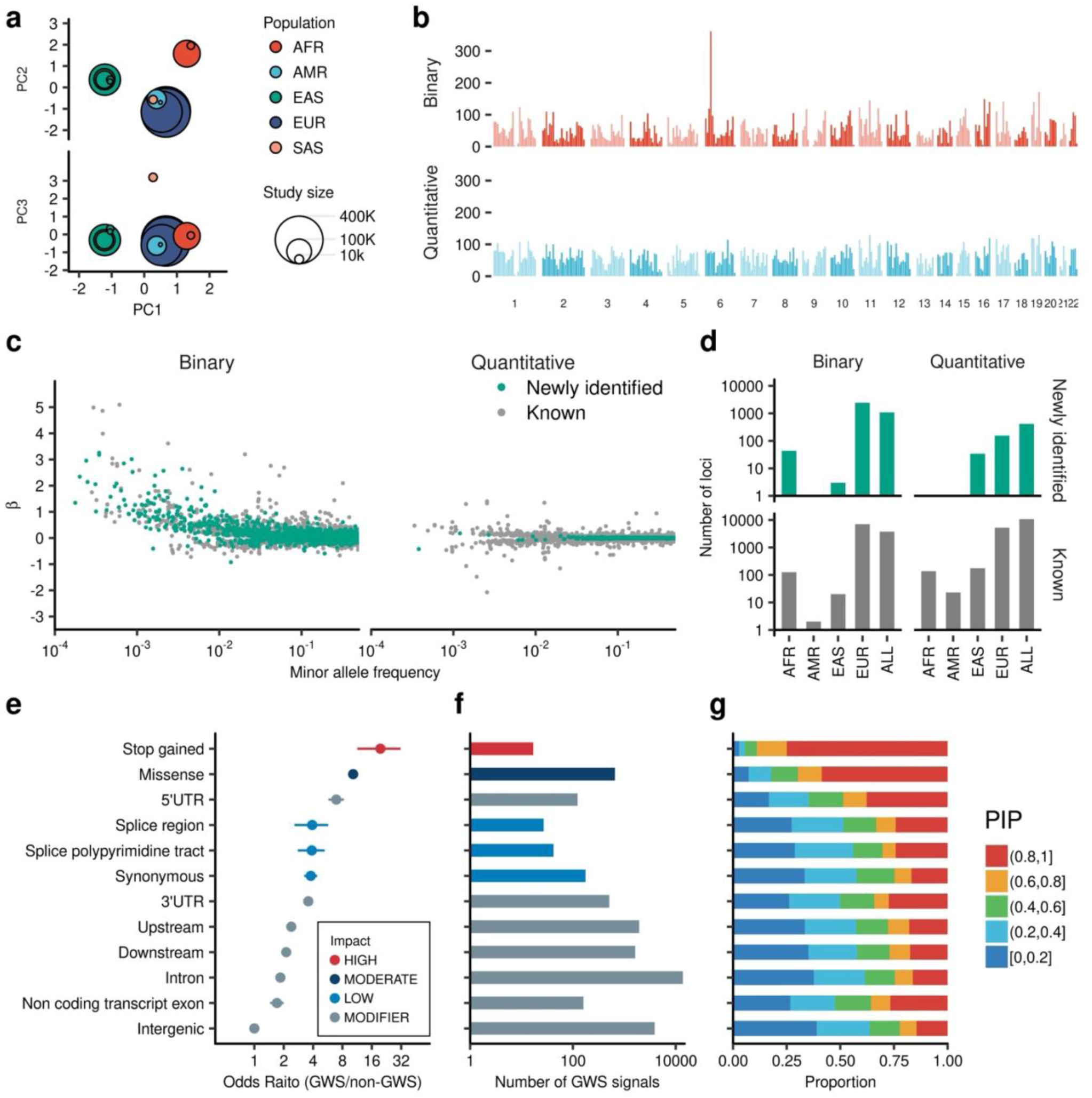
Phenome wide meta-analysis over 1,000 traits. **a**, Overview of the study. The circles show the studies included in this study, by population. Principal components were estimated from MR-MEGA software based on allele frequency of the studies. Circle color indicates genetically similar populations and circle size indicates the number of individuals included. **b,** Genome wide distribution of the associated loci. The height of bars indicates the number of genome wide significant associated loci (non-overlapping 1Mb loci including genome wide significant association with P < 5e-8) in each 10Mb region. Top panel shows the number of loci identified by Binary trait meta-analysis. The bottom panel shows the number of loci identified by quantitative trait analysis. **c,** Effect size distribution of the genome wide significant associations. The horizontal axes indicate minor allele frequency and the vertical axis indicate effect sizes. Each dot indicates trait-variant pairs. The associations that did not reach genome wide significance in the original studies were colored in green. **d,** The number of the loci identified population. **e,** Enrichment of the lead variants in the functional categories. The left panel shows odds ratio for GWS/non-GWS variants in functional categories in reference to intergenic variants. The points indicate estimated odds ratio and error bars indicate its 95% confidence interval. The odds ratios and it’s confidence intervals were estimated by Fisher’s exact test. **f,** The number of the lead variants in each variant category is shown. The color of bars indicates variant categories. **g,** The proportion of each PIP bins by variant functional categories. The horizontal axis shows the proportion of the PIP bins indicted by bar colors. For **e, f,** and **g,** the vertical axes were shared. pLoF, predicted loss of function; UTR, untranslated region; GWS, genome-wide significant; PIP, posterior inclusion probability.

In total, we identified 31,467 non-overlapping locus-trait pairs, driven by at least one genome-wide significant signal in any of the within- or multi-population analyses (307 from AFR, 25 from AMR, 234 from EAS, 14,883 from EUR, and 16,018 multi-population meta-analysis; **Supplementary Figure 1a, Supplementary Table 7**). Among these loci, 13.2% (n = 4,167) did not reach GWS in the contributing studies (**Supplementary Figure 1b, Methods**). Many of these newly identified loci were driven by discovery in the EUR population (3,018/4,167), but multi-population meta-analysis additionally identified 1,029 loci that did not reach GWS in within-population analyses (**Supplementary Figure 2**). For these non-redundant locus-trait associations, we sought replication in the All of Us Research Program (AOU) – an independent cohort spanning the United States (N_Max_ = 206,383). Overall, effect directions were concordant in discovery meta-analysis and replication analysis (97.5% for binary traits, *P* < 1 × 10^−300^ by binomial test and 94.6% for quantitative traits, *P* < 1 × 10^−300^, Supplementary Table 8). The high directional concordance supports the replicability of phecode-based and quantitative phenotyping, genetic associations, and meta-analysis across biobanks (Extended Data Figure 2).

### Coding-variant associations identified by phenome-wide meta-analysis

We identified 86 lead variant-trait associations driven by predicted loss-of-function (pLoF) variants and 1,652 by missense variants. Of these coding variant associations, 7 pLoF associations and 117 missense associations were signals with biologically plausible associations that weren’t identified in the contributing studies. For example, a stop-gain variant (p.Arg259Ter) in the *CLYBL* gene encoding Citramalyl-CoA Lyase was associated with increased risk for “Other Vitamin B12 Deficiency Anemia” (Phecode 281.12, AAF = 2.81%; odds ratio (OR) and 95% confidence interval (95% CI) = 1.21 [1.13-1.29], *P* = 8.85 × 10^−9^). Citramalyl-CoA Lyase is involved in vitamin B12 metabolism with a previously suggested role in Vitamin B12 deficiency anemia;^15^ however, this locus was not identified in prior GWAS at the GWS level. We observed an association between the missense variant *PLAUR* (rs4760, p.Leu317Pro) and increased risk for myocardial infarction (Phecode 411.2, AAF = 15.4%; OR [95% CI] = 1.05 [1.03-1.07]; *P* = 4.52 × 10^−8^). *PLAUR* is the gene encoding the receptor for urokinase plasminogen activator. Plasminogen is an established emergent treatment for acute ST-elevation myocardial infarction when prompt percutaneous coronary intervention is unavailable, and circulating levels of urokinase plasminogen activator receptor have been previously linked to atherosclerosis phenotypes using Mendelian Randomization.^16^ We also observed a novel association between the most common alpha-1-antitrypsin deficiency variant (rs28929474, *SERPINA1* p.Glu366Lys [‘Z-type’], AAF = 1.93%; OR [95% CI] = 1.20 [1.13-1.27]; *P* = 8.15 × 10^−9^) and septicemia. While this variant has been strongly implicated in chronic pulmonary and hepatic diseases, the association with septicemia was not previously observed (OR [95 %CI]_MVP_ = 1.19 [1.11-1.27]; P_MVP_ = 3.20 × 10^−7^; OR [95% CI]_UKBB_ = 1.23 [1.08-1.41]; *P*_UKBB_ = 1.73 × 10^−3^).

Multi-population analyses provide a unique opportunity to identify population-specific associations. One such example is the association between rs624307, an AFR-enriched missense variant in *SLC25A45* (p.Met224Val, MAF_AFR_ = 8.14%, MAF_AMR_ = 0.75%, MAF_EAS_ = 0%, MAF_NFE_ = 0.03%, and MAF_SAS_ = 0.02%) and renal function (estimated glomerular filtration rate, and serum creatinine). *SLC25A45* was previously implicated in renal function through an association with another missense variant (rs34400381, MAF_AFR_ = 0.58%, MAF_FIN_ = 6.46%, MAF_NFE_ = 3.42%) enriched in the Finnish population.^17^ While rs34400381 was significantly associated with renal function in our study, due to the co-existence of another strong signal nearby, rs34400381 was not included in the 95% credible set in our EUR analysis. On the other hand, in the AFR population, rs624307 attained PIP of 1.0 suggesting it may represent a causal variant at this locus. We observed such potentially population specific coding variants in 20 loci in AFR, 1 locus in AMR and 31 loci in the EAS analysis (**Supplementary Table 9**).

### Phenome-wide meta-analysis revealed signatures of genetic associations

To characterize signatures of genetic associations across the phenome, we tested for enrichment of lead variants within genomic annotations. We observed that lead variants were 19.15 times more likely to be stop gain variants (OR [95% CI] 19.15 [11.11 - 30.82]; *P* = 1.8 × 10^−16^; **Figure 1d-e**), and 9.78 times to be missense variants (OR [95% CI] = 9.78 [8.98 - 10.63]; *P* < 1.0 × 10^−300^) compared to intergenic variants (**Figure 1e, 1f**). We also observed a substantial gradient of lead variant enrichment across functional non-coding annotation categories. Namely, 5’UTR variants showed strong enrichment (OR [95% CI] = 6.50 [5.39 – 7.78]; *P* = 3.3 × 10^−56^], with modest but significant enrichment of GWS lead variants in intronic regions relative to intergenic regions (OR [95% CI] = 1.80 [1.74 - 1.86]; *P* = 7.19 × 10^−254^). When we integrated fine-mapping results into our analysis, the posterior inclusion probability (PIP) was highly correlated with variant annotation. We observed a median PIP of 99.8% (interquartile range [IQR]: 96.1 – 100%) for stop gain lead variants (n = 31) and 91.0% (IQR: 49.2 - 1.00) for missense lead variants (n = 657), followed by 5’UTR variants with a median PIP of 61.6% (IQR: 26.5 - 93.3%) and 3’UTR variants with 39.1% (IQR: 17.6 – 75.1%) which were higher than for other annotations (median 27.4% for intergenic variants, n = 3,891, Fig 1g).

To further characterize non-coding lead variants, we explored recently established atlases of regulatory elements developed by the ENCODE consortium, observing that lead non-coding signals were enriched in regulatory elements.^18^ We identified 543, 6,570, and 3,465 lead variants in PLS, ELS, and CTCF binding sites, including 60, 712, and 344 previously unidentified signals, respectively. We observed strong enrichment of putatively causal variants with high PIP in candidate cis-Regulatory Elements (cCREs). The strongest enrichment was observed in promoter-like signatures [PLS, OR = 7.70 (6.49 – 9.07), *P* = 1.66 × 10^−76^], followed by the CCCTC-binding factor (CTCF) binding site [OR = 4.21 (3.88 – 4.56), P = 1.77 × 10^−192^] and enhancer-like signatures [ELS, OR = 3.72 (3.48 – 3.98), *P* = 4.99 × 10^−270^]. One such example is the association between rs146690175 (PIP = 100%) and corneal dystrophy, which was previously unidentified. rs146690175 is located in the PLS, upstream of *NID1* which is known for its role in the formation of the basement membrane.

Tissue specificity of DNA accessibility provides functional insights into the associated variants. Enrichment analyses of the lead variants in tissue-specific DNAse hypersensitivity sites identified 107 significant tissue-trait combinations (35 in binary and 72 in quantitative) with false discovery rate (FDR) < 0.05 (Extended Data Figure 3c, Supplementary Table 10, 11). Among binary traits, we found 11 combinations with binary outcomes and Lymphoid-specific regulatory elements, including hematological cancers (Leukemia, Non-Hodgkin lymphoma) and autoimmune diseases (Asthma, Atopic/contact dermatitis, Hypothyroidism, Psoriasis, and Rheumatoid arthritis). One example is the association between rs72699870 in the intron of *RIN3* gene and Rheumatoid arthritis (OR [95% CI] 0.935 [0.914 - 0.958], *P* = 2.09 × 10^−8^). This variant exists in the active enhancer region in Lymphoid cells and the intronic substitution (ENST00000216487:c.533-1576T>C) disrupts the consensus motif for the YY1 transcription factor. eQTL data suggests this substitution decreases expression level of *RIN3*. These results suggest that trait- and disease-associated genetic variation is often enriched within areas of open chromatin within the specific tissues relevant for the trait/disease.

### Variant-Level Pleiotropy

Systematic genetic analysis including hundreds of traits allowed us to assess pleiotropy at several levels. We first evaluated pleiotropy at the variant level. We defined a pleiotropic variant as a GWS lead variant with multiple significant associations across the wide range of phenotypes we tested (**Supplementary Figure 1c, Supplementary Table 12**). Among 31,467 lead variants identified in the study, 88.38% showed associations with > 1 trait, 27.43% with > 10 traits, and 5.72% in > 30 traits. We observed significant enrichment of pleiotropic variants in functional variants (**Figure 2a**). We found 23 pleiotropic pLoF variants (range 2 – 75 phenotypic associations per variant) and 537 pleiotropic missense variants (range 2 – 86 pleiotropic associations per variant). For example, a splice donor variant in *GCSAML* (rs56043070, MAF 7% in Europeans) showed associations with seven phenotypes, including increased risks of Purpura and other hemorrhagic conditions (PheCode 287), Thrombocytopenia (PheCode 287.3), Urticaria (PheCode 947); decreased Lymphocyte, monocyte, and platelet counts; and increased mean platelet volume. Generally, missense variants showed even greater extents of pleiotropy, such as 68 associations with *APOE* p.Cys130Arg, 65 with *SLC39A8* p.Ala391Thr, 60 with *GCKR* p.Leu446Pro, 51 with *PNPLA3* p.Ile148Met, and 45 with *TM6SF2* p.V727A (**Figure 2b**). *SH2B3* p.Trp262Arg was the most pleiotropic variant identified, broadly associated with 86 binary and quantitative outcomes (**Extended Data Figures 4a and 4b**). *SH2B3* p.Trp262Arg (rs3184504) is known to be associated with various immune-related traits, exemplified by associations with decreased risk of hypothyroidism and systemic lupus erythematosus. The variant allele frequency for *SH2B3* p.Trp262Arg differs widely across populations potentially due to strong selective pressures per recent reporting (**Extended Data Figures 4c** and 4d).^19^ To explore the global relationships between selection signature and phenome-wide pleiotropy suggested by *SH2B3* p.Trp262Arg, we systemically cross-referenced lead variants identified in this study with selection summary statistics^19^. We evaluated changes in allele frequencies of disease-causing alleles in lead variants across different categories of diseases. For example, we found more lead variants whose frequencies decreased in circulatory system (41 decreasing- and 11 increasing-risk alleles; *P* = 3.59 × 10^−5^, **Figure 2c**) and increasing for skin diseases (59 decreasing- and 19 decreasing-risk alleles; *P* = 6.42 × 10^−6^). The variants under selection (selection probability > 20%) were more pleiotropic (**Figure 2d**). For example, variants with selection probability > 80% were 6.45 times more likely to be pleiotropic (OR [95% CI] = 6.09 [3.99 – 9.78], *P* < 1.78 × 10^−29^). Furthermore, selection probability is positively associated with the number of pleiotropies (**Figure 2e**). For example, 24.2% (295/1,219) of variants with more than 30 pleiotropies showed a selection probability greater than 80%, compared to 0.77% (22/2,857) of non-pleiotropic variants. Finally, we observed that highly pleiotropic variants showed stronger selection coefficient. One such example is rs8176693 which is under positive selection (S = 2.94%, SE = 0.3%, *P* = 8.2 × 10^−23^) and associated with 31 traits. Taken together, variants influencing a greater number of phenotypes are more likely to be targets of selection.

**Figure 2.**
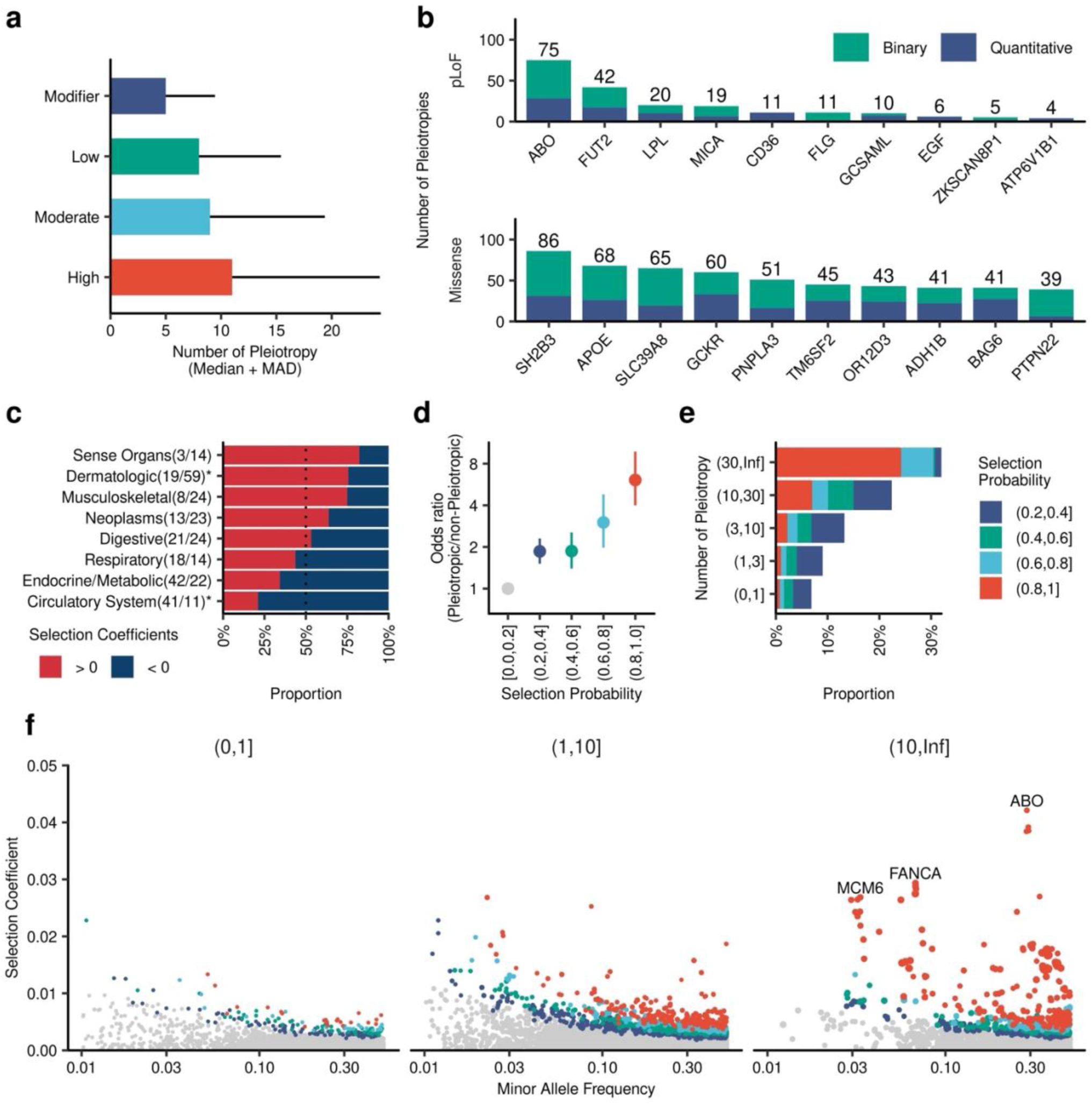
Variant Level Pleiotropy. **a**, Median pleiotropy by variant classes. The bar charts show the median number of pleiotropic phenotypes by variant class. The error bars represent mean absolute deviation. Star (*) Indicates a significant difference tested by the Wilcoxon rank-sum test (P < 0.05/8). **b,** Top 10 pleiotropic genes with pleiotropic coding variants. The numbers on top of the bar charts indicate the number of pleiotropies. **c,** Selection signatures by PheCodes categories. Red and blue colors indicate the number of GWS lead variants (*P* < 5 × 10^−8^) with high posterior probability for selection (> 0.99) to increase and decrease disease prevalence (Red: selection to increase the risk allele; Blue: selection to decrease the risk allele). **d,** The enrichment of pleiotropic variants under selection pressure. The enrichment of variants with multiple GWS associations in variants with specified selection probability in reference to posterior probability for selection (0-0.2). The error bars show the 95% confidence interval of odds ratio estimated by Fisher’s exact test. **e,** The enrichment of variants under selection in pleiotropic variants. The color of bar charts shows the posterior probability for selection. **f,** Larger selection coefficients in pleiotropic variants. The horizontal axes show minor allele frequency, and the vertical axis shows the selection coefficient (under positive or negative selection selection). pLoF, predicted loss of function; GWS, Genome wide significant.

### Locus-level Pleiotropy

We next evaluated the extent of pleiotropy at the level of physical genomic loci. In aggregate, trait-associated variation fell within 2,624 non-overlapping autosomal genomic windows. Each window with at least one association contained GWS variant-trait associations for a median of 6 traits (IQR 2 to 18), including 2,110 (80%) pleiotropic regions containing an association with two or more traits.^20^ To distinguish groups of traits sharing true genetic signals rather than signals confounded by linkage disequilibrium, we performed multi-trait colocalization.^20,21^ Overall, high-confidence (posterior probability > 0.9) evidence of colocalization was observed at 72% (1,902/2,624) of genomic regions, forming 3,687 clusters of traits in EUR, 520 in AFR, 502 in EAS, and 162 in AMR (**Supplementary Table 13**). Although these high-confidence pleiotropic clusters occurred throughout the genome, this distribution was not equal across chromosomes (Fisher’s exact test p < 0.001), consistent with prior reports (**Supplementary Figure 4**).^4^ Rather, the distribution of pleiotropic clusters across the genome was proportional to chromosome length (Spearman R = 0.85, p = 1 × 10^−6^) and the number of genes per chromosome (Spearman R = 0.89, p = 3.3 × 10^−6^).

Next, we classified each pleiotropic cluster based on the phenotypic categories represented by their constituent traits. In aggregate, domain-specific pleiotropy (e.g. a shared signal across several phenotypes within a single phenotypic category) predominated, accounting for 74% of all pleiotropic clusters. However, there was significant variability in the proportion of domain-specific signals observed across population subgroups, ranging from 70% in the EUR population to 90% within the AFR subgroup (Fisher’s exact test *P* < 0.001) (**Figure 3b**). Although multi-domain signals were less common, putative causal variants at multi-domain signals were significantly enriched in several functional annotations when compared to putative causal variants at domain-specific signals, including coding variants, genomic regions conserved across species, and regulatory elements, among others (**Figure 3c; Supplementary Table 14**).

**Figure 3.**
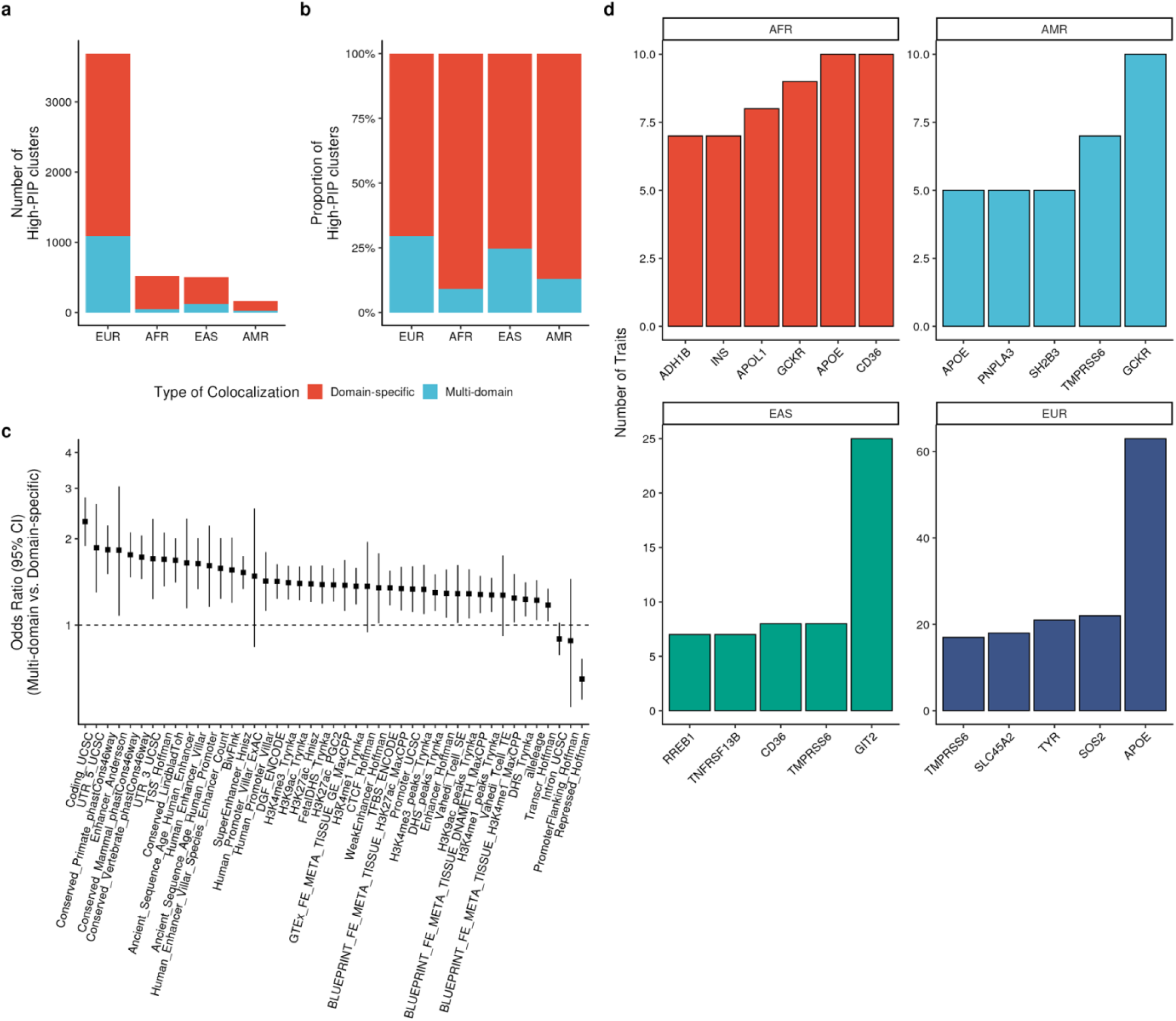
Locus-level pleiotropy. **a**, Number of signals with high posterior inclusion probability for colocalization (PIP > 0.9) across 2+ traits within each population, with colors representing whether clusters were domain-specific (including phenotypes within a single category), or multi-domain (phenotypes spanning multiple categories). **b,** Proportion of domain-specific and multi-domain signals across population subgroups. **c,** Enrichment of lead variants at pleiotropic loci in functional annotations, comparing multi-domain to domain-specific signals. **d,** Loci where coding variants link multiple traits with strong evidence of colocalization (PIP > 0.9).

Given the strong enrichment of coding variation in pleiotropic loci, we evaluated the patterns of pleiotropy at loci where coding variants linked multiple traits with strong evidence for colocalization (PIP > 0.9). Among EUR, the pleiotropic coding signals associated with the greatest number of phenotypes occurred at the *APOE* locus, which encodes apolipoprotein E, a key component of circulating lipoproteins including low-density lipoprotein (LDL). At the *APOE* locus, the two missense variants (rs429358 and rs7412) that contribute to the APOE E2/3/4 alleles influenced 63 traits across 5 different phenotype categories, including established links to risk of Alzheimer’s Disease and several cardiometabolic traits. Among AFR, the most pleiotropic signal occurred at the *CD36* locus, yielding associations for 10 traits (**Figure 3D**). *CD36* encodes a membrane glycoprotein on a wide variety of cell types.^23^ The candidate causal variant at this locus (rs3211938) encodes the *CD36* p.Tyr325Ter stop-gain variant. This variant is significantly more common among AFR than other populations in gnomAD (AF_AFR_ = 0.09 vs. AF_Other_ = 0.00039, Fisher’s exact *P* < 0.001). It has previously been reported to be under positive selection in AFR populations, likely due to its link to malaria resistance.^24,25^ A large pleiotropic cluster at *CD36* (rs75326924; p.Pro90Ser) influencing 8 traits was also present among EAS. However, the largest cluster of signals among EAS occurred at the *GIT2* locus, where the missense variant rs925368 (p.Asn387Ser) was associated with 25 traits including many blood cell traits and cardiometabolic traits/diseases. Finally, the largest cluster among AMR occurred at the *GCKR* locus, where the missense variant rs1260326 (p.Leu446Arg) was associated with 10 traits related to circulating lipids, obesity, and renal and hepatic function.

Because most pleiotropic signals were non-coding, we further tested whether they were enriched in more highly-constrained regions of the genome as defined by the Gnocchi score.^22^ Both multi-domain and domain-specific signals were significantly enriched in the most constrained non-coding regions of the genome (**Supplementary Table 15**). Our results suggest that pleiotropic signals and those affecting multiple phenotype categories are under selection and enriched across important functional domains.

### Global Pleiotropy

To characterize the genetic architecture of traits and diseases at the global level, we estimated genome-wide measures of population-specific heritability and polygenicity. Compared to data from a recently published analysis of the VA Million Veteran Program, we found that our meta-analysis identified significant heritability (h^2^ Z > 4) for more traits across a broader spectrum of populations (**Supplementary Table 16**). Heritability estimates were strongly correlated across populations (Spearman R 0.66 to 0.93, *P* < 0.001 for all pairwise comparisons), and traits with population-specific effective sample sizes over 15,000 had heritability estimates that were even more strongly correlated between population groups (R > 0.80 between all population group pairs, *P* < 1.04 × 10^−11^) (**Supplementary Figure 5**). Cross-population genetic correlations were also high, with medians ranging from p_gi_ 0.61 to 0.83 in pairwise analyses (**Supplementary Table 17, Supplementary Figure 6**).^26^ Polygenicity estimates (proportion of causal variants) ranged from 2.67 × 10^−5^ to 0.027 and the distribution of polygenicity was similar across populations (a median of approximately 0.5% of variants were estimated be causal per phenotype; **Supplementary Figure 7, Methods**). However, consistent with prior reports, trait-level polygenicity estimates varied across populations (**Supplementary Table 18**), likely due to differences in selective pressures and adaptation.^27^

Bivariate genetic correlations were estimated for pairs of traits within each population (**Supplemental Table 19**). For several highly heritable (h^2^ Z > 4) traits, we observed scenarios where no significant genome-wide correlation was detected between traits despite strong evidence of locus-specific colocalization driven by coding variation (**Supplementary Table 20, Supplementary Figure 8**), consistent with differences in the global and locus-level architectures of complex traits.

### Genetic prediction (Polygenic Scores)

We developed multi-population polygenic scores (PGS; n = 1,167) using PRS-CSx, a method that leverages coupled continuous shrinkage priors to model variant effects across diverse populations. Scores were developed using GWAS summary statistics from our population-specific meta-analyses, and performance was evaluated among participants of AOU with available genotype and phenotype information. We observed that models including a PGS significantly (FDR < 0.05) improved upon baseline models containing only age, sex, and genetic principal components for the majority of phenotypic endpoints (73.9% [95% CI 70.6 to 76.9%; 568/769] among AFR; 100% [95% CI 99.6 to 100%; 1,216/1,216 phenotypes] among EUR, **Supplementary Table 21**). These significant incremental associations were driven by 265 unique PGS among AFR, and 435 PGS among EUR, representing 22.7% (95% CI 20.4 to 25.2%) and 37.3% (95% CI 34.5 to 40.1%) of all newly constructed PGS (n = 1,167), respectively.

To determine how our new multi-population PGS performed in comparison to existing risk scores, for each Phecode-based outcome we compared predictive performance against existing scores from the Polygenic Score (PGS) Catalog. There were PGS for n = 514 phenotypes among AFR, and n = 1,216 phenotypes among EUR where both our newly constructed multi-population PGS and previously published PGS Catalog Scores significantly (FDR < 0.05) improved upon the baseline model (age, sex and genetic principal components). Among AFR, the incremental R^2^ was greater using the newly constructed PGS for 97% (500/514; 95% CI 95.3 to 98.4%) of phenotypes, and among EUR the incremental R^2^ was greater using the newly-constructed PGS for 65.5% (797/1,216; 95% CI 62.8 to 68.2%) of phenotypes (**Supplementary Figure 8**). These findings demonstrate that our newly constructed PGS substantially improve upon the predictive performance of a large breadth of existing PGS, particularly among the AFR population.

Finally, we performed phenome-wide scans in AoU, testing associations between each PGS and all clinical endpoints. We observed high levels of pleiotropic/“off-target” associations. Among EUR participants, each PGS significantly (FDR < 0.05) improved upon a baseline model for a median of 121 endpoints (IQR 45 to 249), while among AFR participants each PGS improved upon a baseline model for a median of 3 endpoints (IQR 1 to 10). For example, among EUR the most pleiotropic PGS included non-specific/symptom-related scores like “Nonspecific chest pain” (Phecode 418; 709 associated endpoints), “Shortness of breath” (Phecode 512.7; 686 associated endpoints), and “Pain in limb” (Phecode 773; 682 associated endpoints). Among AFR participants, the most pleiotropic PGS included BMI (137 associated endpoints), weight (128 associated endpoints), and “Obesity” (Phecode 278.1; 127 associated endpoints). We also observed target phenotypes that were predicted by large numbers of PGS. Among EUR participants, a median of 82.5 PGS (IQR 26 to 236) associated with each endpoint, and among AFR participants a median of 4 PGS (IQR 2 to 13) associated with each target endpoint. The endpoints predicted by the most PGS among EUR participants were “Tobacco use disorder” (Phecode 318; 889 associated PGS), “Mood disorders” (Phecode 296; 853 associated PGS), and “Substance and addiction disorders” (Phecode 316; 833 associated PGS). Among AFR participants the endpoints predicted by the most PGS included “Type II diabetes” (Phecode 250.2; 141 associated PGS), “Tobacco use disorder” (Phecode 318; 137 associated PGS), and “Diabetes mellitus” (Phecode 250; 126 associated PGS). Overall, these results suggest that the specificity of PGS varies considerably, with each PGS often associated with multiple endpoints, and endpoints often predicted by multiple PGS.

## DISCUSSION

In genome-wide analyses of 1,167 traits spanning the phenotypic spectrum among 1,633,308 participants of diverse biobanks, we developed a comprehensive catalog of variant-phenotype associations. By combining data from six major biobank initiatives and validating findings in the All of Us Research Program, we identified 31,467 genome-wide significant locus-trait pairs, including 4,167 associations that had not reached genome-wide significance in previous phenome-wide GWAS efforts. Systematic analysis of these genetic associations revealed pervasive pleiotropy at the variant, locus, and trait levels, advancing our understanding of the shared genetic basis of human traits and diseases while demonstrating the value of diverse population representation in genomic studies. Our findings have important implications for future large-scale human genetics research, both for biological discovery and disease prediction.

Our GWAS findings, including 4,167 newly-discovered locus-trait associations, demonstrate the value of increasing both sample size and genetic diversity of populations included in large-scale human genetic studies, consistent with prior reports for individual phenotypes and biobanks.^6,28,29^ The identification of previously undiscovered associations, particularly those specific to non-European populations, underscores the importance of diverse population representation in genomic research. For example, the discovery of the AFR-enriched missense variant in *SLC25A45* associated with renal function highlights how population-specific analyses can reveal putative causal variants that might be missed in studies of European populations alone. Similarly, the observation of different selection pressures across disease categories - negative selection for cardiovascular diseases and positive selection for dermatologic diseases - suggests how distinct evolutionary forces may have shaped disease risk across human populations in the face of contemporary exposures. The enrichment of variants under selection pressure in pleiotropic regions indicates that pleiotropy may play a crucial role in both human adaptation and disease susceptibility.

The extensive pleiotropy observed at multiple levels - variant, locus, and trait - provides insights into the shared genetic architecture of human traits and diseases. Our observation that 72% of loci demonstrate evidence of pleiotropy, in contrast to 84% of lead GWAS variants, suggests that some pleiotropic associations are driven by “apparent” (eg. confounded by linkage disequilibrium) rather than “true” pleiotropy; however, most GWAS loci do appear to be shared across two or more phenotypes, consistent with prior reports.^4^ Importantly, we observe that loci influencing phenotypes across different phenotypic categories are enriched for important functional annotations, suggesting these loci may represent more critical biological processes that are shared across broad categories of traits and diseases. We also find that at many loci coding variation is responsible for pleiotropic associations, and these pleiotropic coding associations frequently span multiple categories of traits/diseases. These observations have important implications for future therapeutic development – whether these highly pleiotropic loci affecting broad categories of traits and diseases represent a set of “core” targets that could be targeted to prevent/treat many diseases, or conversely represent targets likely to have important undesired consequences will require future study.

The development and validation of multi-population polygenic scores represents an important advance in genetic risk prediction. The enhanced performance of our newly-constructed PGS compared to existing scores, particularly among the AFR population of All Of Us, reinforces the value of incorporating broad populations in genetic studies. This improvement in predictive performance has the potential to help improve risk stratification and personalized medicine strategies across populations. Importantly, we observe that individual PGS often have associations with a large number of “off-target” phenotypes, and each phenotype is often predicted by dozens of PGS developed primarily for other traits. While recent PGS methods have begun to leverage “off-target” associations to further improve genetically-informed risk models of disease^3,30^, this high degree of PGS pleiotropy suggests that PGS-based risk models should be interpreted cautiously, as PGS associations may be driven by broad sharing of genetic architecture, rather than representing precise/phenotype-specific genetic associations.

### Limitations

Several important limitations should be considered when interpreting our findings. First, despite our efforts to include biobanks studying individuals with varying genetic backgrounds, European population participants still predominate our analysis (EUR, N_Max_ = 1,282,152 versus AFR, N_Max_ = 128,206), potentially limiting our ability to detect population-specific associations and affecting the generalizability of our findings to non-European populations. Second, the differing disease prevalences across biobanks, stemming from differences in recruitment strategies and healthcare system patient-populations could influence differences in power and estimated effect size estimates. Third, while our phenome-wide approach captures a broad spectrum of traits, the reliance on EHR-derived phenotypes may introduce misclassification bias, particularly for conditions that are challenging to diagnose or require specialist care. Fourth, our analysis of pleiotropy, while comprehensive, may still underestimate the true extent of shared genetic architecture due to limited statistical power for rarer variants and traits. Fifth, the cross-sectional nature of most biobank data limits our ability to assess temporal relationships and disease progression, which may influence patterns of pleiotropy. Finally, while our multi-population polygenic scores show improved performance, the continued population-based disparities in predictive accuracy highlight the ongoing need for more diverse genetic studies. Future research with more balanced population representation, longitudinal follow-up, and harmonized phenotyping approaches across biobanks will be valuable in addressing these limitations.

### Conclusion

Our findings emphasize the importance of considering pleiotropic effects in genetic studies, while highlighting the value of broad population representation in genomic research. This resource will facilitate the investigation of shared genetic architecture across traits toward mechanistic insights and therapeutic development as well as the construction of more accurate genetic prediction tools across populations.

## METHODS

### Ethics oversights

All data utilized for this manuscript was obtained from publicly available resources. The underlying biobanks were overseen by their respective institutional review boards, and all participants provided informed consent.

### GWAS meta-analysis

All summary statistics were publicly available and obtained from each cohort on the following dates: September 2, 2023, for BBJ (https://pheweb.jp/); December 8, 2023, for FINNGEN (https://www.finngen.fi/en); August 7, 2023, for KOGES (https://koges.leelabsg.org/); August 8, 2023, for UKBB (https://pan.ukbb.broadinstitute.org/); July 20, 2024, for MVP (https://ftp.ncbi.nlm.nih.gov/dbgap/studies/phs002453/phs002453.v1.p1/analyses/GIA/); and August 22, 2024, for TMM (https://jmorp.megabank.tohoku.ac.jp/gwas-studies). First, we processed all the summary statistics on the hg38 reference genome, using the liftOverVcf function implemented in GATK as needed. Then, we computed LD-score intercept using LDSC and multiplied standard errors by the root of LD-score intercept to have LD-score intercept of 1 for all the summary statistics. We filtered variants with minor allele counts of < 10 before meta-analysis. Traits were combined across biobanks using a combination of manual mapping for quantitative traits, and computational harmonization of phecodes/ICD codes. Where possible, phecodes were directly mapped across cohorts using the Phecode map v1.2 (http://phewascatalog.org). When cohorts reported trait-specific associations using ICD-codes, harmonization to phecodes was performed using Jaccard similarity, as previously-described.^31^ Population-specific GWAS meta-analysis for each trait was then performed separately for AFR, AMR, EAS, and EUR populations using GWAMA. For multi-population analysis, we conducted meta-regression using MR-MEGA, which uses genetic principal components to further account for population stratification and partition genetic associations correlated with allele frequency variation/population. Only variants observed more than 5 studies were considered in MR-MEGA analyses to allow for calculation of genetic principal components based on allele frequencies in each population. In multi-population analyses, effect sizes (beta and standard error of beta) were calculated using GWAMA, while statistical significance was determined by P-values computed by MR-MEGA. A conventional GWAS p-value threshold of 5×10^−8^ was used to identify regions of the genome with putative associations with traits/diseases for further downstream analyses of pleiotropy. This p-value threshold was selected because 1) control of test-statistic inflation was performed using LD score regression (see below) minimizing false-positive associations; 2) external replication of all significant findings was pursued in the All of Us research program, and the empiric false-positive rate was assessed for well-powered traits in the replication analysis; 3) Downstream analyses considering enrichment, colocalization, genetic correlation, and polygenic risk scores were performed to validate the genetic architecture of all traits at the variant, locus, and global levels, including several techniques that do not depend on arbitrary p-value thresholds (eg. LD-score regression, colocalization, and Bayesian polygenic risk scores.

### LD score regression

We computed the LD-score intercept using the ldscr R package for summary statistics from the original study, utilizing population-matched reference LD-scores provided by the Pan-UKBB project. To control inflated test statistics, we multiplied the standard error of the original summary statistics by the squared LD-score intercept. This correction adjusts the LD-score intercept to be 1 before meta-analysis. We further computed the attenuation ratio for summary statistics from within-population meta-analysis. The attenuation ratio was calculated as

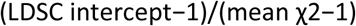

Here, the attenuation ratio is the measure of non-polygenic inflation if the lambda GC was high, as previously described.^32^ LD score regression was also performed to estimate population-specific heritability on the observed scale, and to estimate bivariate genetic correlations between pairs of heritable (h^2^ z-score > 4) traits as previously recommended.^33^

### Definition of trait-associated loci

We determined trait-associated loci using physical position. First, we extracted all the variants with genome wide significant associations (P < 5×10^−8^) and added +/-500Kb flanking regions to each range. Overlapping ranges were then merged using the *bedtools merge* function. This process generates non-overlapping regions with a minimum length of 1Mb. To identify newly-associated loci compared to the input datasets, we defined previously identified associations based on the original summary statistics used in the meta-analysis. After performing within-population and multi-population meta-analyses (Supplementary Tables 5 and 6), we merged all loci and selected the strongest signal with the smallest P-value (Supplementary Figure 1a, Supplementary Table 7). The 31,467 merged loci and 23,107 unique lead variants were used in the downstream analysis.

### Variant annotation

We used VEP version 107 to annotate variants. To assign a single annotation for each variant, we used the “--pick” option implemented in VEP which prioritize deleterious annotation in the high confidence transcript.

### Replication

We queried all the lead variants identified in population-specific and multi-population meta-analysis from pre-computed GWAS summary statistics available from the All Of Us Research Program. Details of genotyping and association testing are available at https://allbyall.researchallofus.org/. We assessed the concordance of the population-wise associations by populations (AFR, AMR, EAS, and EUR). The multi-population metanalysis was compared with fixed-effect meta-analysis results in the AOU. We stratified associations based on statistical power, which was calculated using the effect size from the meta-analysis, effect allele frequency, and the number of cases and controls in AoU.

### Finemapping

We computed the posterior inclusion probability (PIP) under the assumption of a single causal variant. PIP was calculated for each GWS locus in both within-population and multi-population meta-analyses. For the fixed-effect meta-analysis, we used the following formula for Bayes factor finemapping:^28^

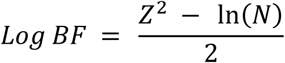

For the multi-population meta-analysis, we used the log Bayes factor computed by MR-MEGA. To characterize the relationship between PIP, variant annotation, and other downstream analyses, we took the highest PIP among phenotype– variant pairs to represent their potential to be causal variants.

### Pleiotropic Variants

To assess the extent of pleiotropy at the variant level, we queried 23,107 lead variants across all meta-analysis summary statistics. If the P-values were less than 5 × 10⁻⁸, we considered the variant pleiotropic for those traits. While querying all summary statistics (4 intra-population analyses and 1 multi-population analysis), if a variant was significant in multiple populations, we counted it as a single pleiotropy.

### Pleiotropic loci and colocalization

To define pleiotropic loci, lead trait-associated genetic variants were clustered into independent genomic windows, merging variants located with 250kb, and extending each locus +/-100kb upstream and downstream. Regions containing associations for >1 trait were considered pleiotropic. To more rigorously define pleiotropic signals within physically-overlapping loci, multi-trait colocalization was performed using *HyPrColoc*.^21^ This Bayesian statistical approach evaluates the probability that two or more traits share the same signal of genetic association within a given region, accounting for the possibility that multiple groups of traits sharing independent variants may exist within the same region (eg. Traits 1 and 2 share variant A, while traits 3 and 4 share variant B, all within the same genomic region). Within each genomic region, we identified GWAS traits containing at least 1 genome-wide significant association. For regions containing evidence of genome-wide significant signals for two or more traits, colocalization was performed using the default regional and alignment priors: *P*_*R*_ ∗= *P*_*A*_ ∗= 0.5, and high-probability colocalization was defined based on a posterior probability of colocalization of 0.9 consistent with prior reports.^4^ Candidate causal variants within each colocalization cluster were defined based on fine-mapping under single causal variant assumption.^21^ Because the goal of the colocalization analyses was to distinguish true pleiotropy from apparent pleiotropy on the basis of linkage disequilibrium, multi-trait colocalization was performed separately by population subgroup to account for linkage disequilibrium differences due to genetic background.

Pleiotropic signals were classified as “domain-specific” or “multi-domain” based on the phenotypic categories of traits contributing to each cluster. To define domains, GWAS phenotypes were mapped to pheCode categories.^34^ Clusters containing traits from a single category were considered “domain-specific”, while clusters containing traits from multiple categories were considered “multi-domain.”

### Enrichment of functional annotations

Functional consequences of genomic regions were identified using publicly available .bed files provided by the authors of LDSC.^35^ Enrichment of variation within these categories between sets of genetic variants was evaluated using Fisher’s exact test. Enrichment in regions of non-coding constraint was performed using Fisher’s exact test as previously described.^22^

### Polygenicity

Population-specific polygenicity estimates were calculated as previously described.^36^ Briefly, for each heritable trait HapMap3 variants were merged with the corresponding population-specific LD scores generated from UK Biobank participants.^29^ Variants were then grouped into LD score bins, and within each LD score bin adaptive shrinkage regression was performed using the ‘ashr’ package in R to identify the proportion of variants null and non-null effect sizes.^37^ Finally the ‘nls’ package in R was used to fit the relationship between LD score bin and the proportion of non-null variants, with the exponential rate parameter serving as an estimate of polygenicity.

### Polygenic Scores

Multi-population polygenic score weights were generated for each trait using PRS-CSx, using population-specific LD reference panels derived from 1000 Genomes Phase 3 Reference panel.^38^ SNP effects combined across populations using inverse-variance weighting by specifying the “--meta” flag, and the global shrinkage parameter was learned by specifying the “--auto” flag. PGS weights were then applied to genotype information in the All of Us research program using pgsc_calc. Because PGS performance may vary due to differences in population structure, score mean and variance were normalized using the external 1000 Genomes + HGDP reference panel (the Z_norm2 method in pgsc_calc).^39,40^ Associations were tested between each PGS and each phecode-based clinical endpoint with 200 or more cases using a phenome-wide association study (PheWAS) approach using generalized linear models adjusted for age, sex, and the first ten genetic principal components. Models including PGS were compared to baseline models including only covariates using likelihood ratio tests. PGS performance was assessed separately among EUR and AFR AoU participants. To account for multiple comparisons in the context of non-independent PGS and non-independent clinical endpoints, the false discovery rate was controlled at 5% using Benjamini Hochberg to establish PGS that significantly improve model fit compared to the baseline model. Existing scores from the PGS Catalog (accessed August 2024) were calculated using the same approach, and incremental r^2^ (compared with baseline) for PGS Catalog Scores was subtracted from the incremental r^2^ of the newly-constructed scores to identify the proportion of phenotypes with better incremental predictive performance using the newly-constructed scores.^39^

## Data Availability

Genetic association data are available at https://platlas.cels.anl.gov

## Code Availability

The analyses reported in this manuscript rely on previously published software, as detailed in Methods.

## Acknowledgements

The authors thank the participants of the contributing biobanks. M.G. Levin was supported by the Doris Duke Foundation (Award 2023-0224) and US Department of Veterans Affairs Biomedical Research and Development Award IK2-BX006551. This publication does not represent the views of the Department of Veterans Affairs or the United States Government. S. Koyama was supported by NHLBI K99HL169733. D.Y. Zhang was supported by NHLBI F30HL172382. B.F. Voight was supported by NIDDK UM1DK126194. P. Natarajan was supported by NHLBI R01HL127564.

---

**Extended Data Figure 1.**
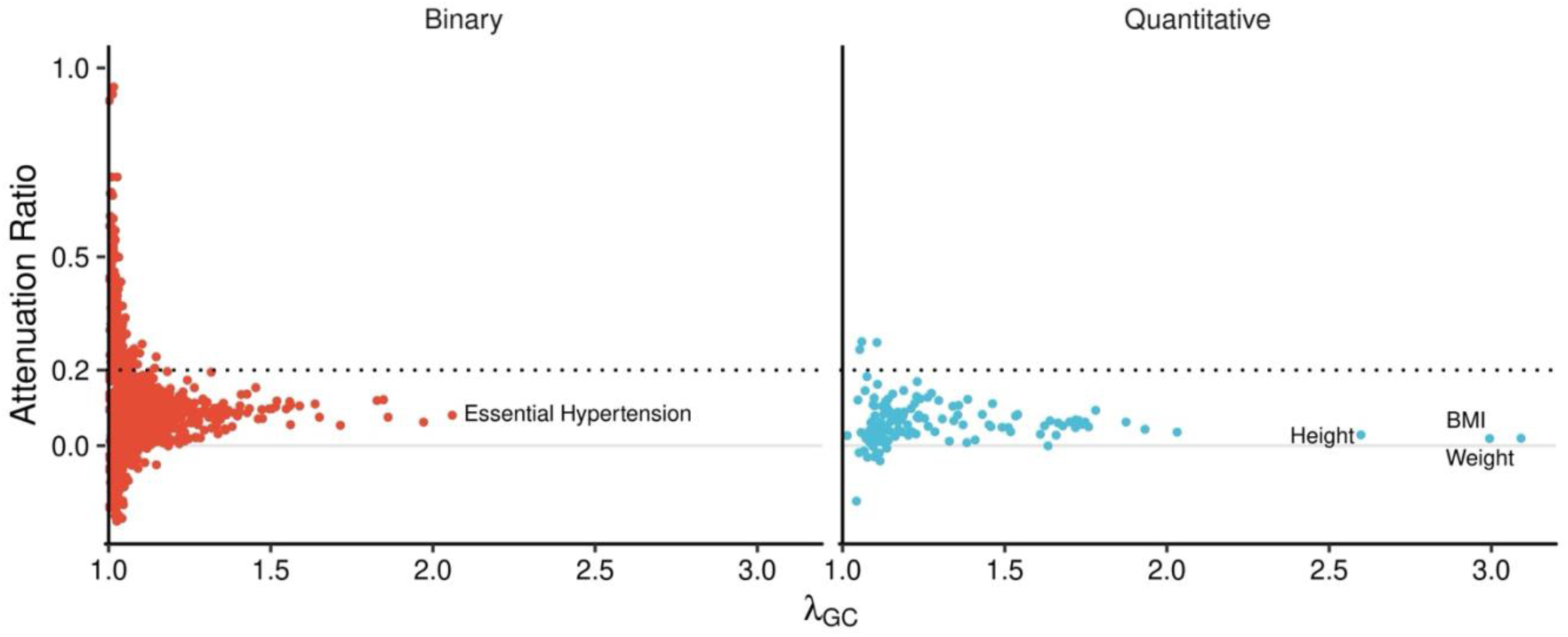
Inflation statistics for within-population meta-analysis. Each dot indicates within-population meta-analysis. The horizontal axes show lambda GC values and the vertical axis shows attenuation ratio. Attenuation ratio was computed by (LDSC intercept – 1)/(mean χ2 – 1). Dotted lines indicate the level of attenuation ratio of 0.2. BMI, Body mass index.

**Extended Data Figure 2.**
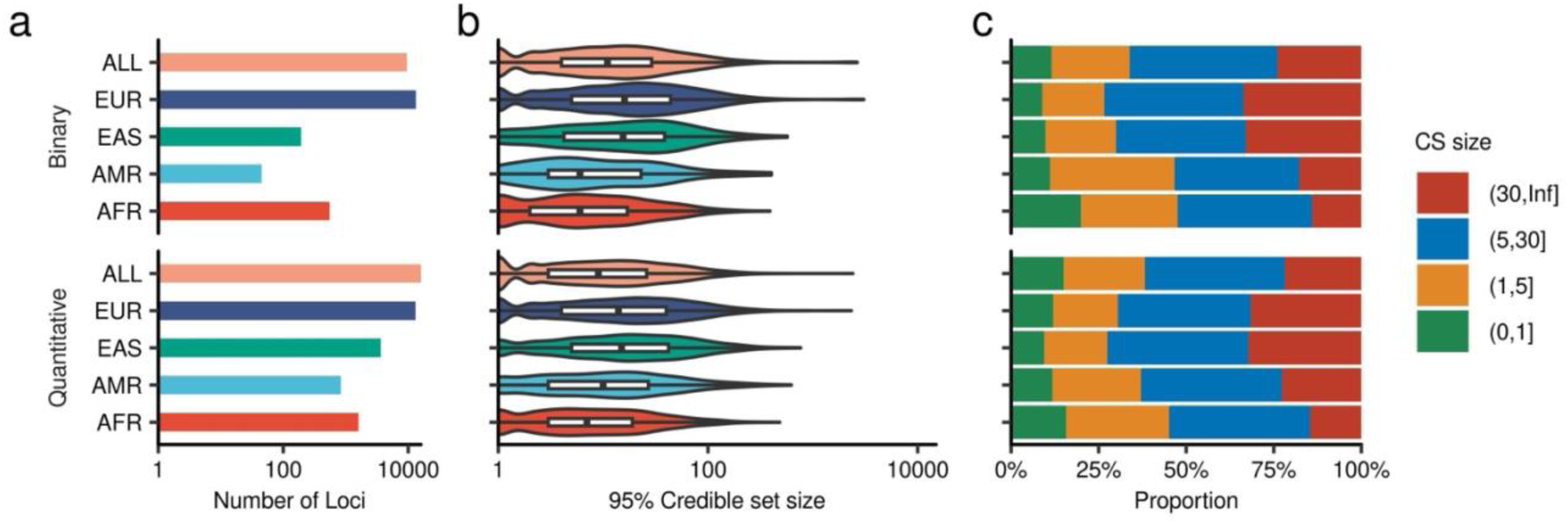
Phenome Wide Genetic Finemapping. The left panel shows the number of loci identified within-population (AFR, AMR, EAS, EUR) and multi-population meta-analysis. The middle panel shows the distribution of the size of 95% credible set sizes (the number of variants in 95% credible set). The right panel shows the proportion of credible set sizes. CS, credible set; AFR, African-like population, AMR Admixed-American-like population, EAS, East-Asian-like population; European-like population.

**Extended Data Figure 3.**
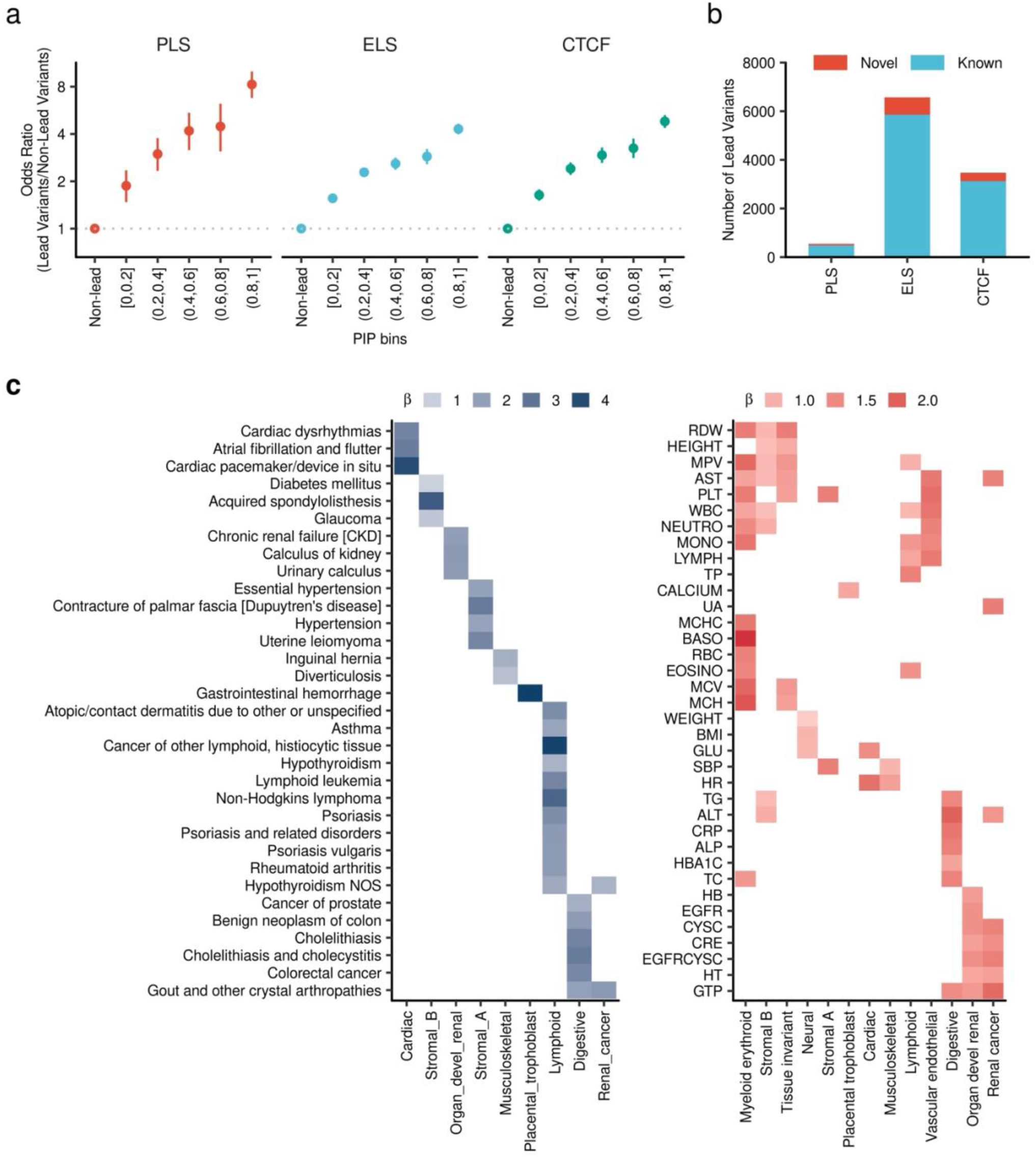
Lead variant Enrichment in the tissue specific regulatory elements. **a**, Enrichment of GWS lead variants in candidate cis-regulatory elements (cCRE). The horizontal axis shows PIP bins and the vertical axis shows odds ratio of the designated PIP bins relative to Non-lead variants. The error bars shows 95% confidence intervals. **b,** Number of lead variants reside in the cCRE. **c,** The heat map shows the enrichment beta (log odds ratio) of lead variant in each trait in the tissue specific DNase hypersensitivity site (Meuleman et al. Nature 2020). The significant combinations between traits and DNase hypersensitivity site are shown (FDR < 0.05). The odds ratios and P-values were computed by Fisher’s Exact Test.

**Extended Data Figure 4.**
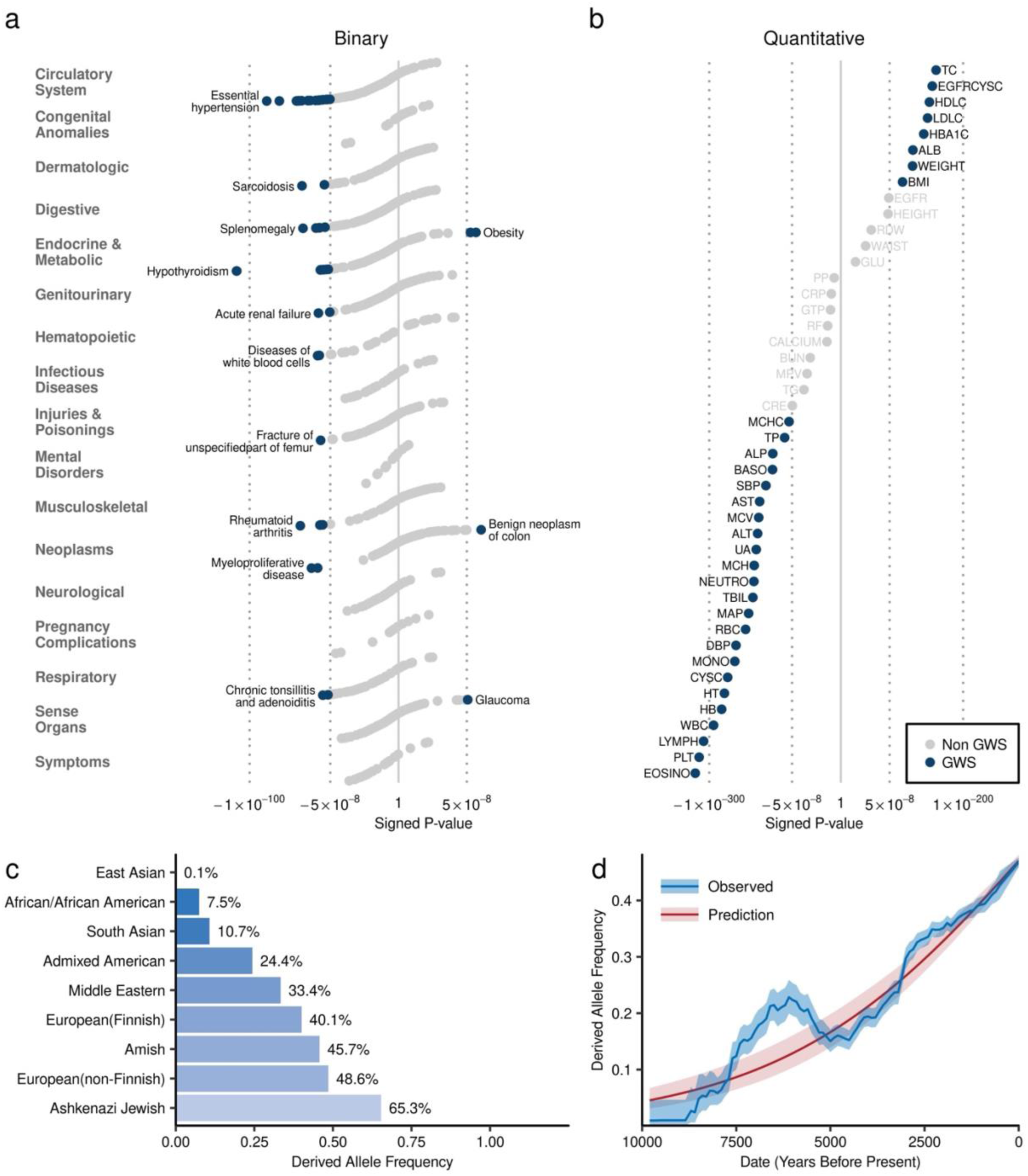
A pleiotropic variant – SH2B3 p.Trp262Arg (rs3184504) **a**, Phenome-wide association of SH2B3 p.Trp262Arg (rs3184504). The dots indicate the P-values for associations between SH2B3 p.Trp262Arg and phenome-wide binary outcomes. The horizontal axis shows the signed negative log10 P-values. **b,** Phenome-wide association of SH2B3 p.Trp262Arg with quantitative traits. Blue dots indicate genome-wide significant associations (P < 5 × 10^−8^). **c,** Allele frequencies of SH2B3 p.Trp262Arg in various populations. The height of each bar indicates population-specific allele frequencies based on the gnomAD dataset. **d,** Allele frequency trajectories of SH2B3 p.Trp262Arg in populations of European ancestry. The horizontal axis represents years before present. The vertical axis shows the estimated allele frequency of SH2B3 p.Trp262Arg at the designated time points. The blue curve and corresponding blue shaded area indicate the estimated allele frequency and its 95% confidence interval. The red curve and corresponding pink shaded area indicate smoothed estimates and their 95% confidence intervals. GWS, genome wide significant.

**Supplementary Figure 1:**
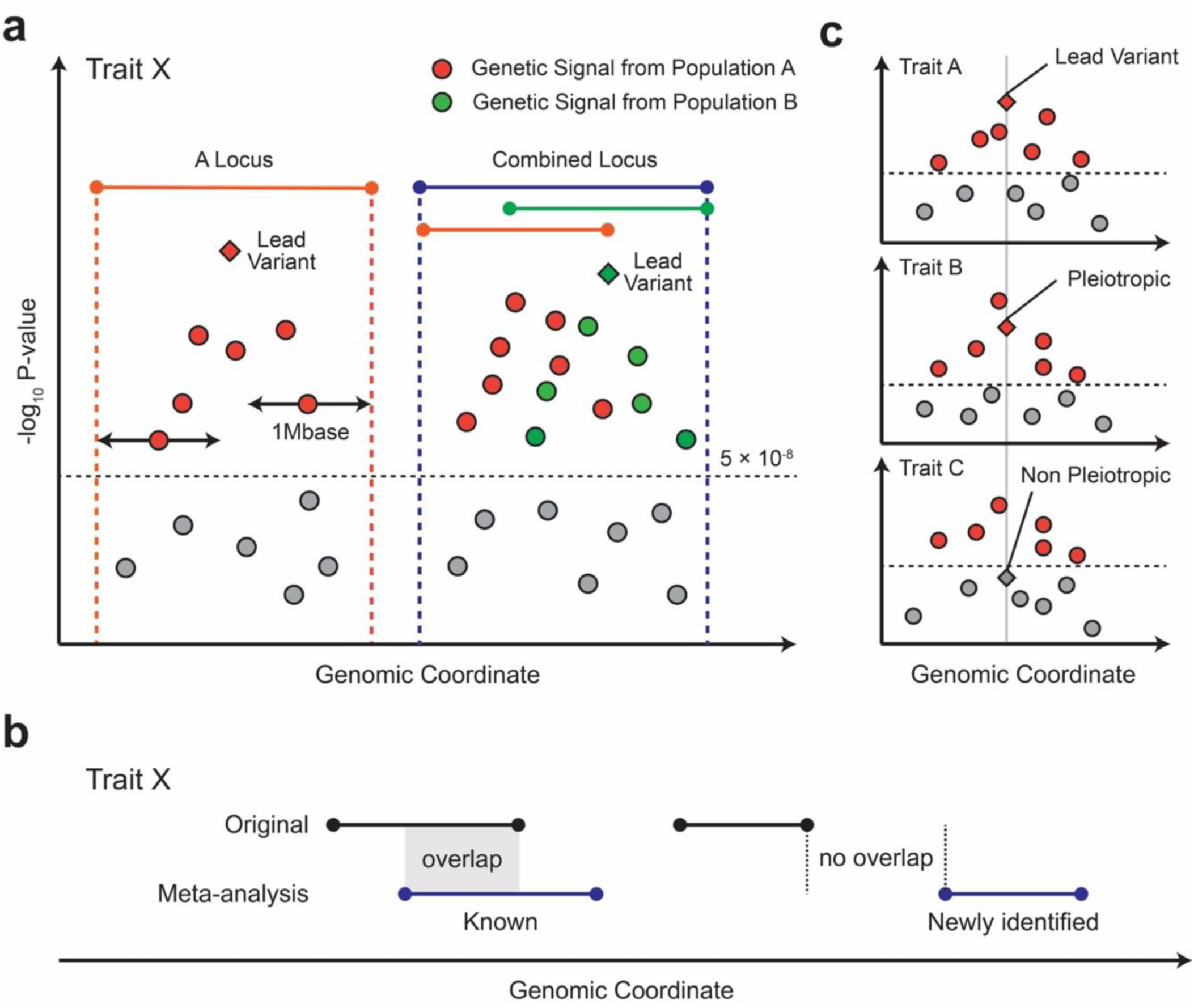
Definition of a locus, newly identified locus, and variant level pleiotropy. a. The locus was defined by physical position on the genomic coordinate. We added 500kb flanking for all the variants with genome wide significant threshold and merged these regions to define a locus with a genome wide significant locus. This locus has at least 1Mb long and not overlapping with the other loci in the same analysis. b, Novelty was considered using overlap with the loci in the original (pre-metanalysis) summary statistics. If the locus was overlapping any of the loci which attained genome wide significance in the original summary statistics that locus was considered as known, and if there is no overlapping with other loci, that locus was annotated as newly identified.

**Supplementary Figure 2.**
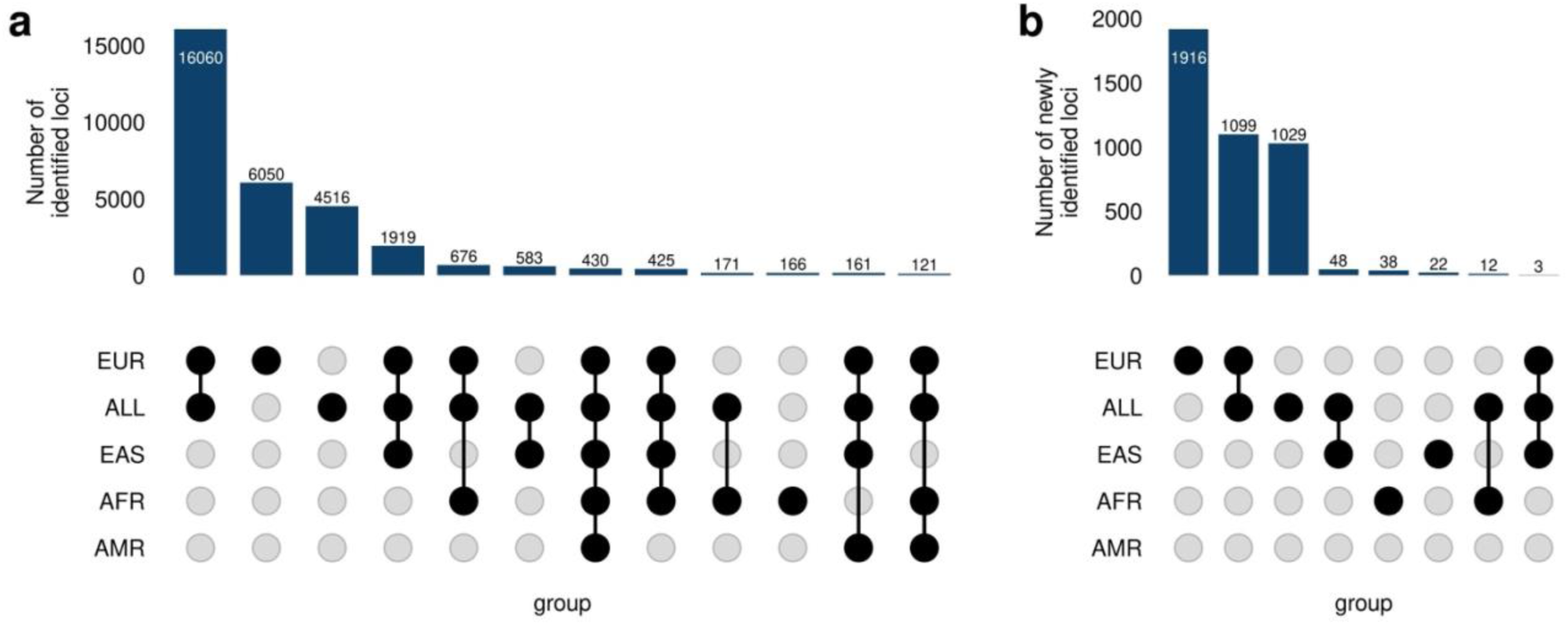
Genome-wide significant signals across populations. **a**, Total numbers of GWS loci by the analysis. The upset plot shows the combinations

**Supplementary Figure 3.**
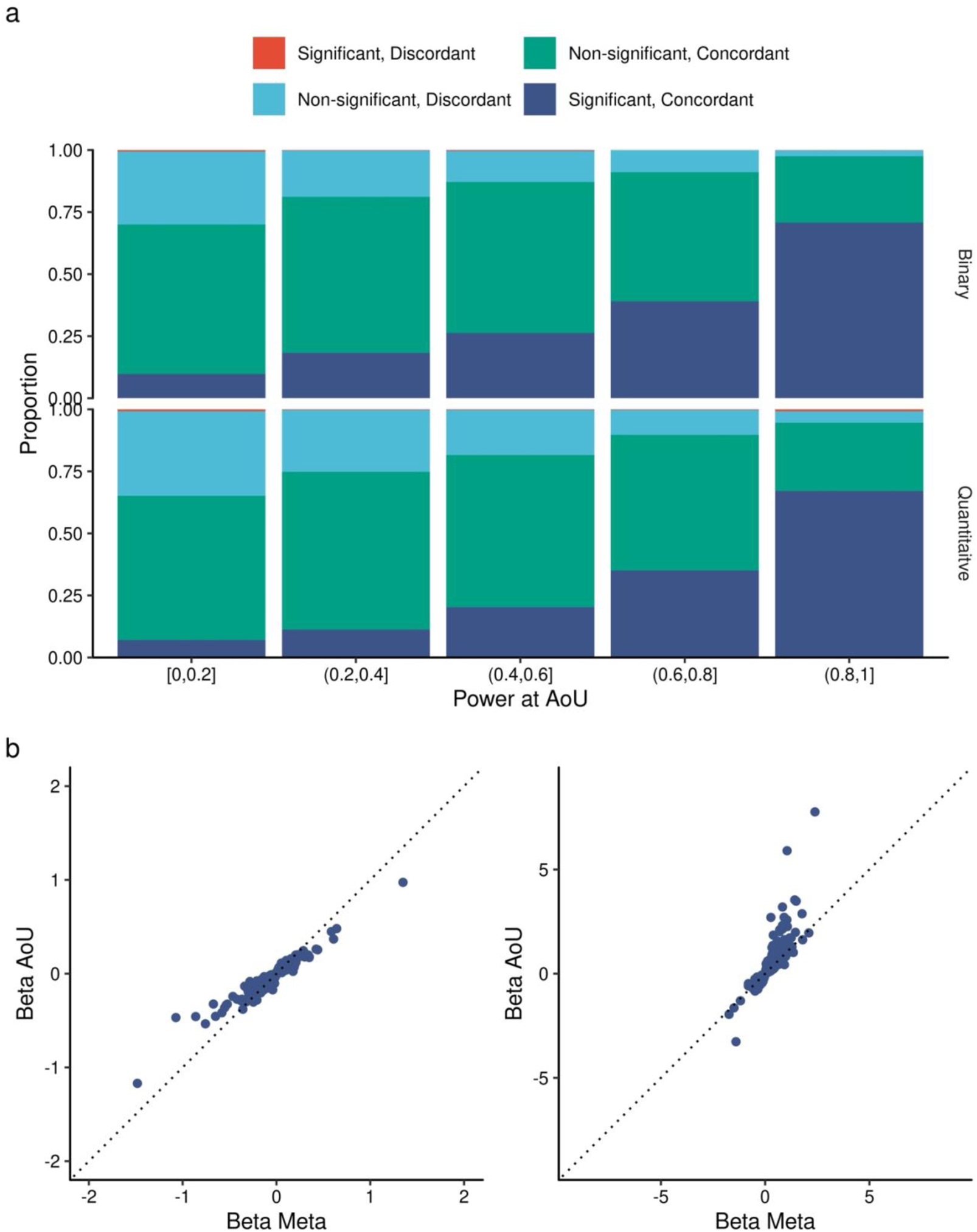
Replication of the association signals. **a**, Replicability of the effect directions of associated signals between meta-analysis and the independent AOU cohort. The colors of the stacked bar charts indicate the proportions of association categories: 1) direction discordant and nominally significant (P_AOU_ < 0.05), 2) direction discordant and non-significant (P_AOU_ ≥ 0.05), 3) direction concordant and non-significant (P_AOU_ ≥ 0.05), 4) direction concordant and significant (P_AOU_ < 0.05). The data is presented separately by the statistical power of detecting associations at P < 0.05 in the AoU cohort. **b,** Effect size comparison between meta-analysis and AOU. The horizontal axis shows the effect size in Meta analysis, and vertical axis shows the effect size in AOU. The left panel shows binary traits, and the right panel shows quantitative traits. AOU, All Of Us project.

**Supplementary Figure 4.**
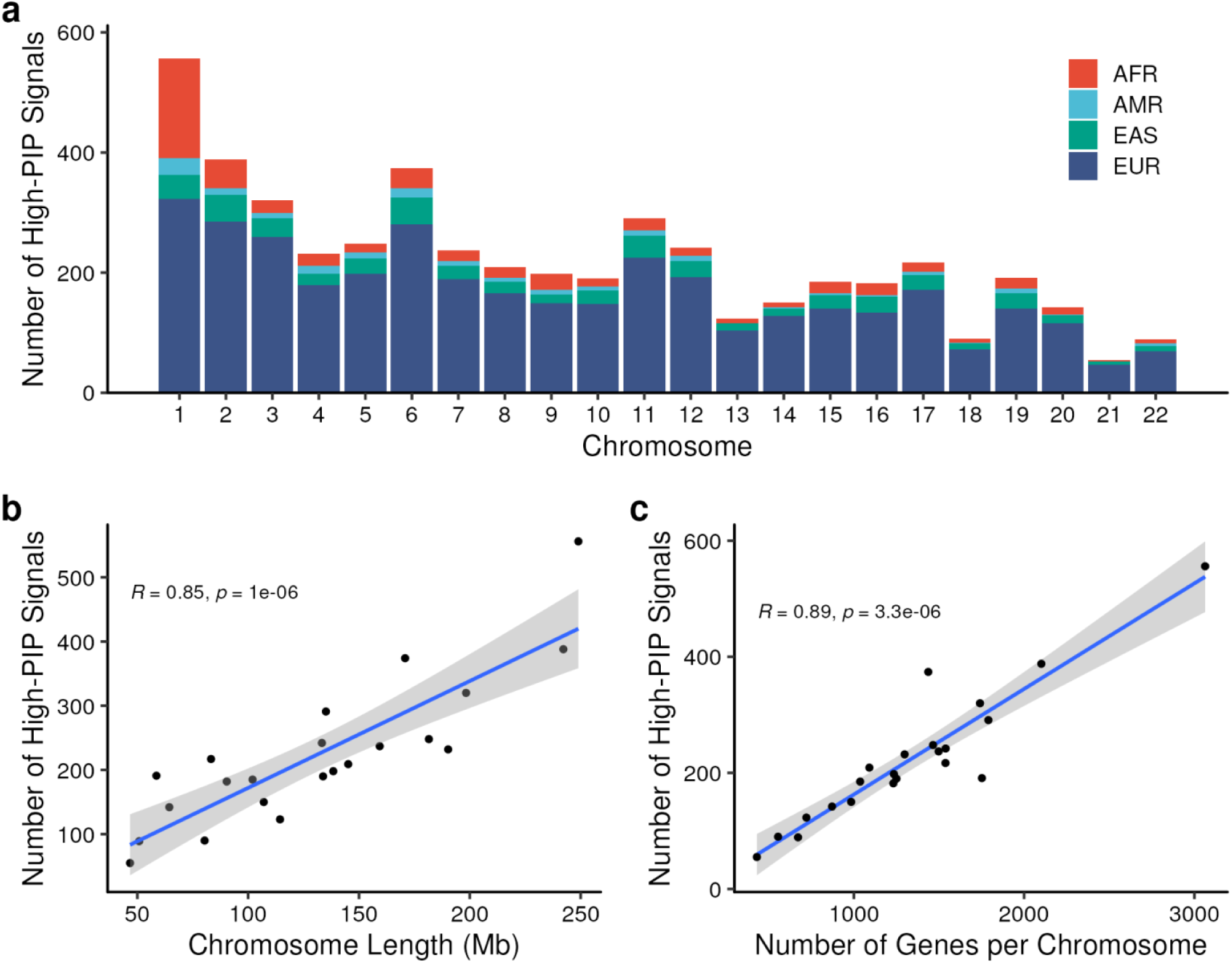
Distribution of Pleiotropic Signals. **a**, Number of signals with high posterior inclusion probability for colocalization (PIP > 0.9) for 2+ traits with each population subgroup (AFR, AMR, EAS, EUR). **b**, Correlation between chromosome length and number of high-PIP signals. **c,** correlation between number of genes per chromosome and number of high-PIP signals.

**Supplementary Figure 5.**
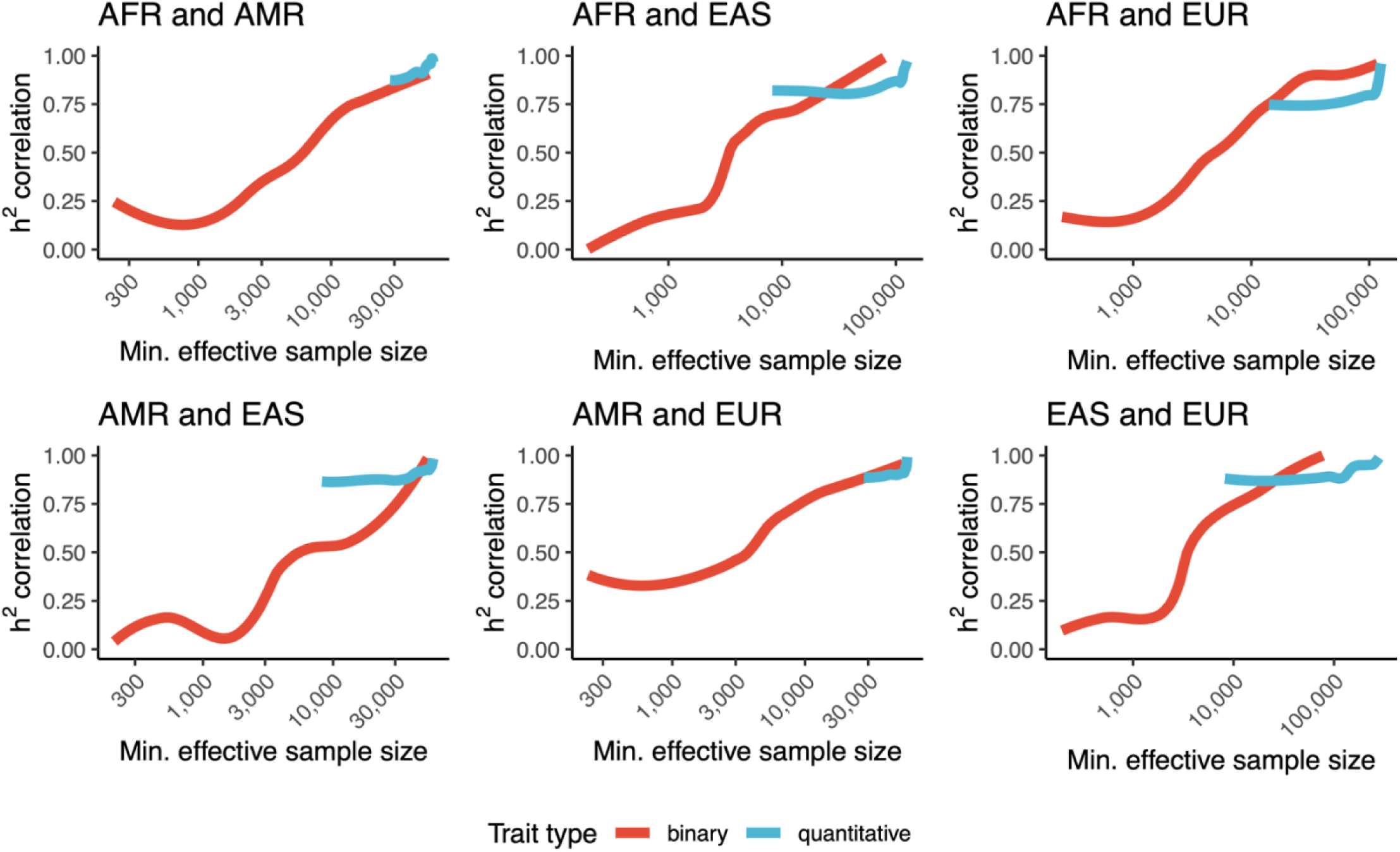
Heritability Correlation across Populations. Pearson correlation of heritability estimates for all pairs of GWAS in common between all pairs of populations. X-axis represents the minimum effective sample size between the pair of population-specific summary statistics on a log scale. The correlation includes all traits above a certain threshold established by the X-axis. Traits are stratified by outcome type, with quantitative traits using sample size, and binary traits using effective sample size computed according to the METAL method. Curves were smoothed with span = 0.5. Includes population-stratified meta-analyzed GWASs and GWASs only in UKB or MVP.

**Supplementary Figure 6.**
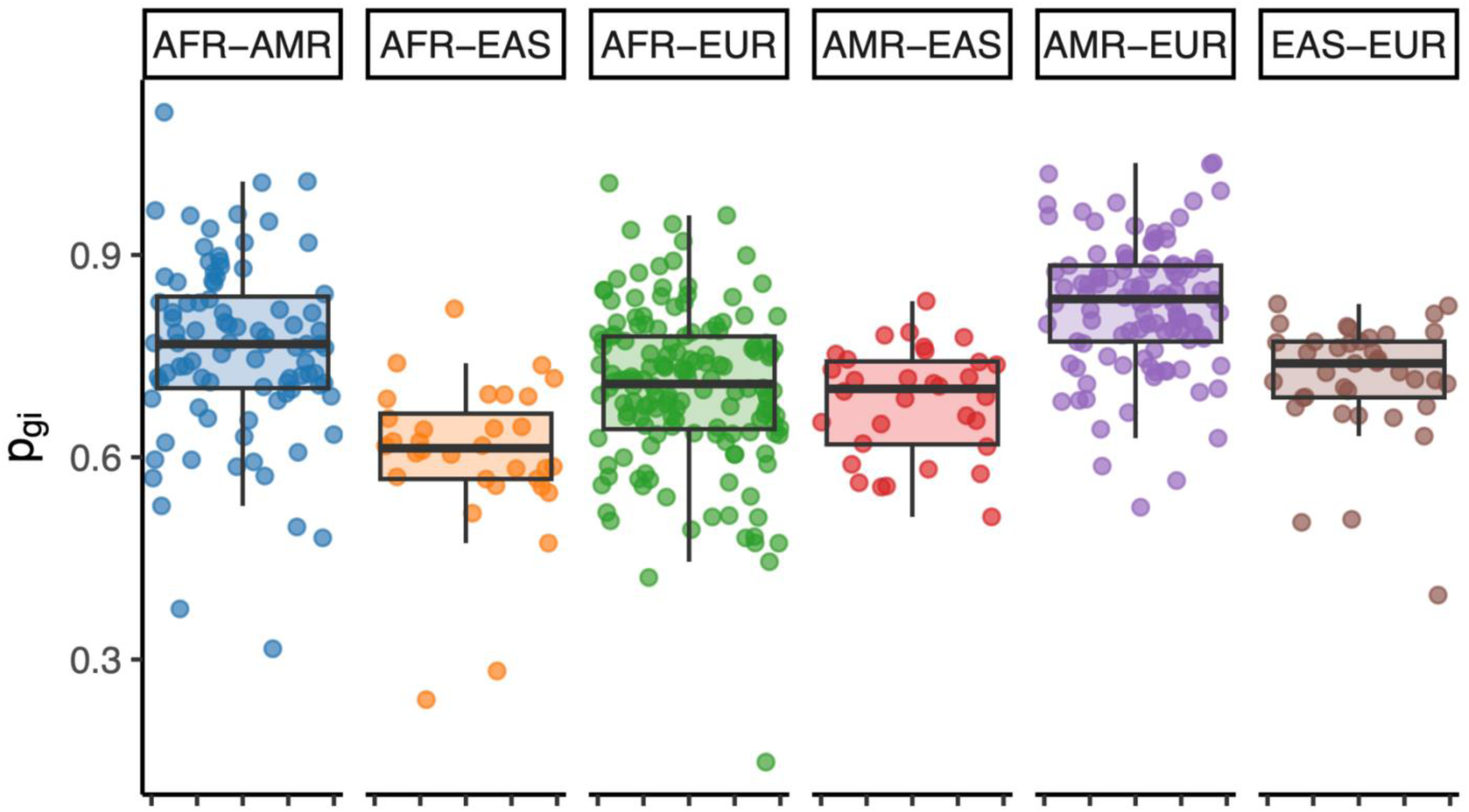
Cross-population genetic correlations. Cross-population genetic correlation estimates (p_gi_) from Popcorn, computed between all pairs of heritable (Z > 4) GWAS and between each population group. Traits were considered heritable if they had significant LDSC h^2^ estimates in both populations.

**Supplementary Figure 7.**
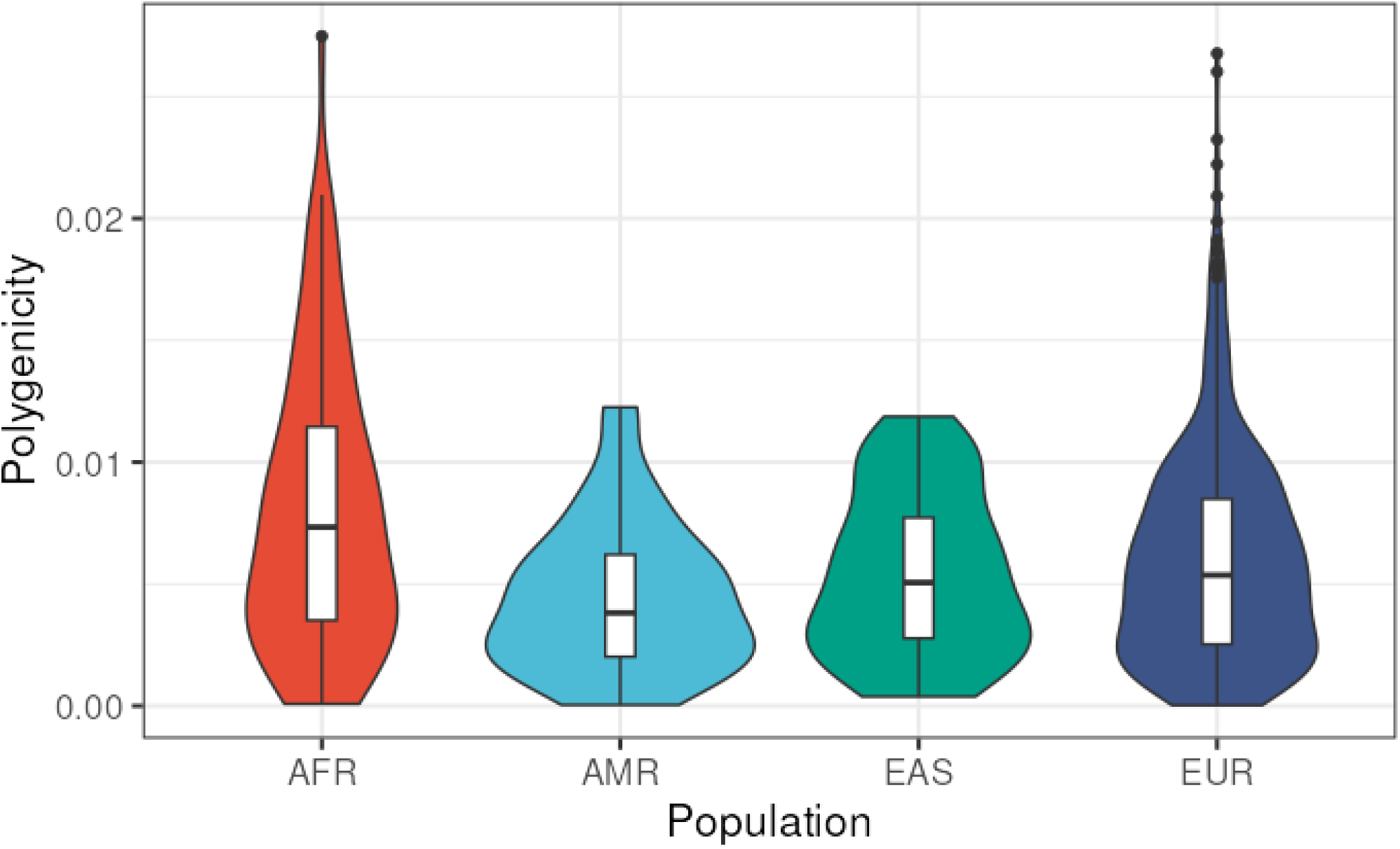
Polygenicity Estimates. Polygenicity was estimated for highly heritably (h2 Z-score > 4) using adaptive shrinkage regression. Polygenicity of traits were similar across populations.

**Supplementary Figure 8.**
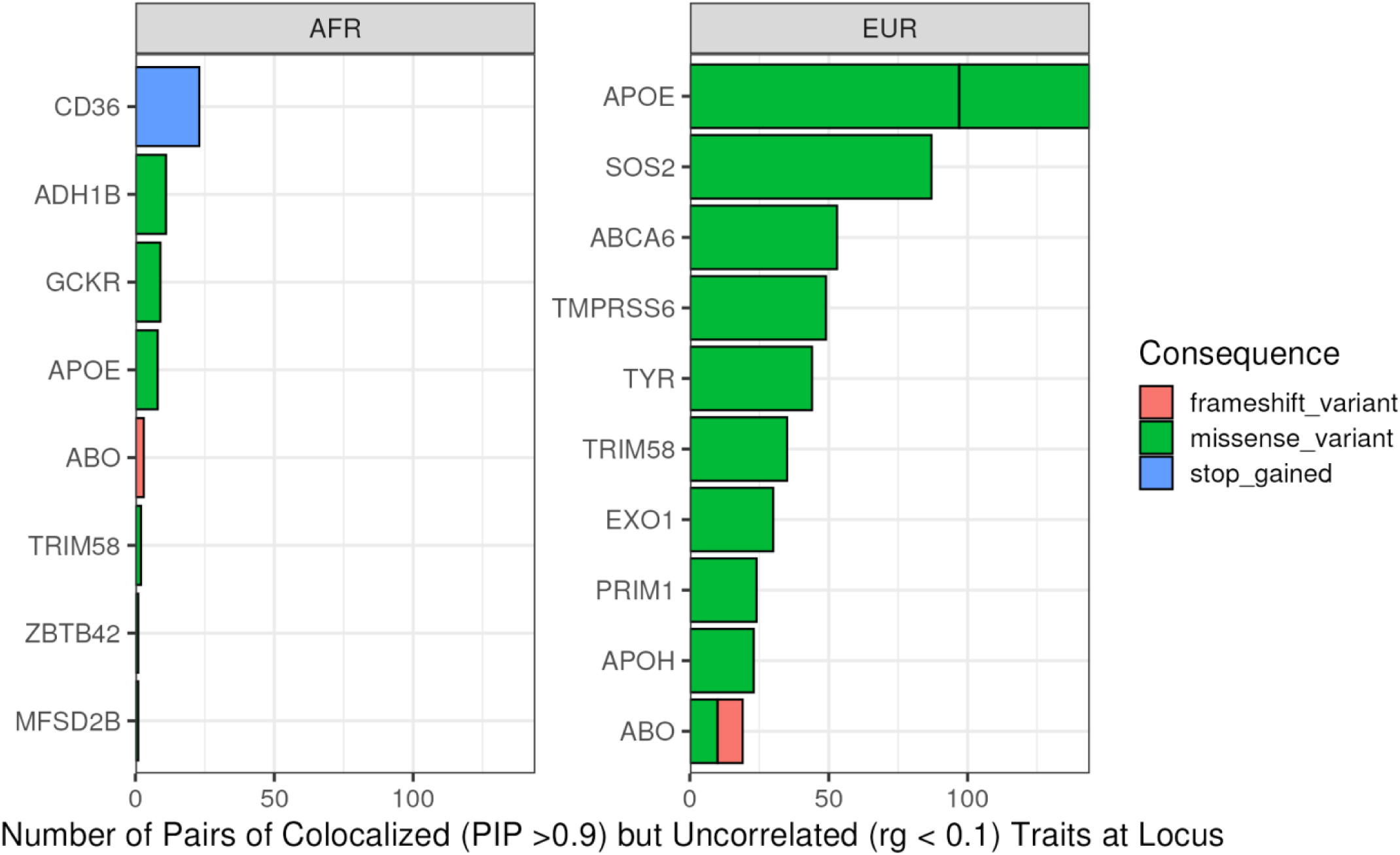
Colocalized Traits without Significant Global Genetic Correlation. Top loci where globally uncorrelated (genetic correlation [rg] < 0.1) traits were linked by strongly-colocalized (Posterior Inclusion Probability [PIP] > 0.9) loci, driven by coding variants.

**Supplementary Figure 9.**
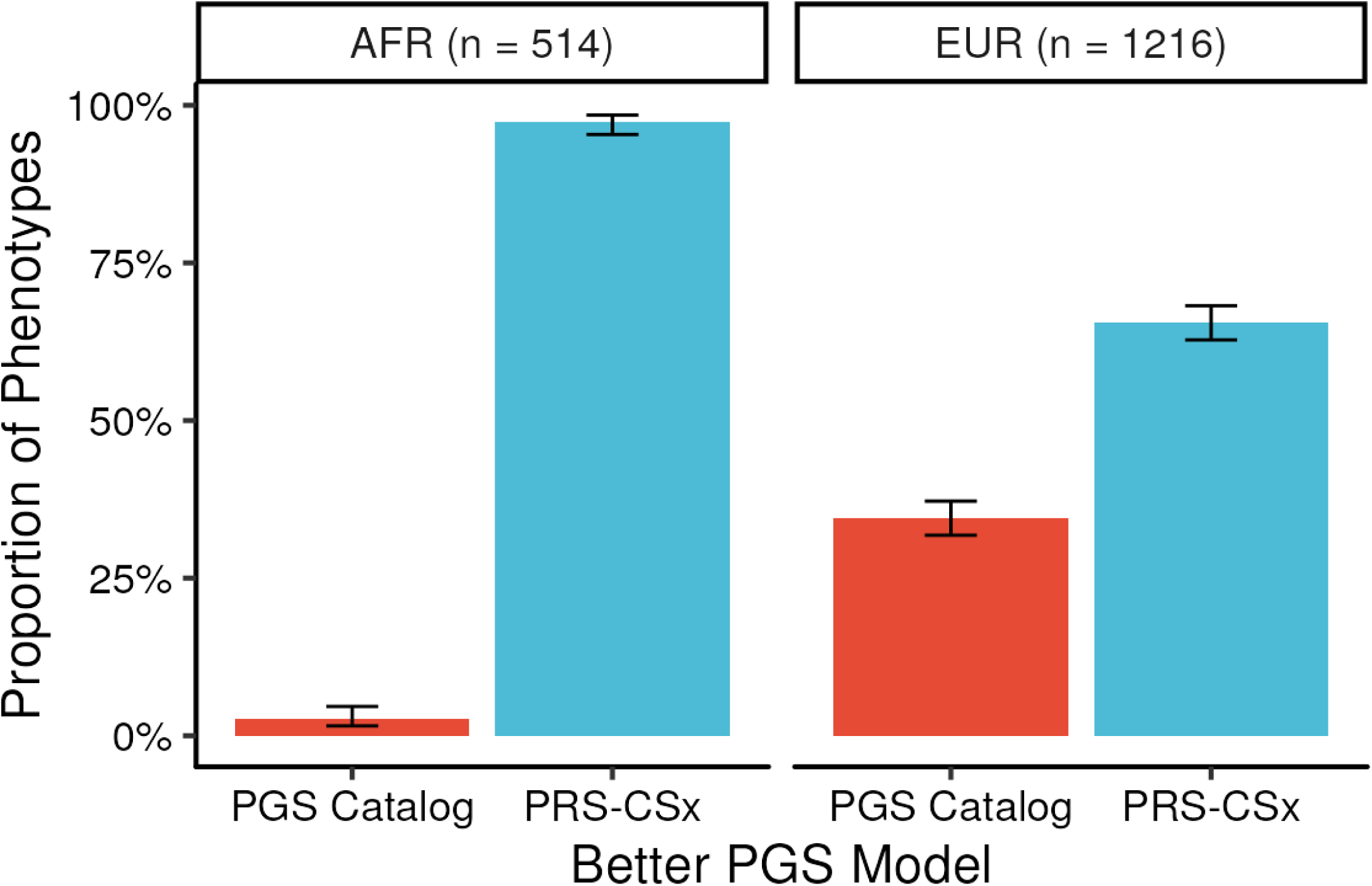
Polygenic Score Comparison. Among phenotypes where polygenic scores from both the PGS Catalog and PRS-CSx significantly improved upon a baseline model (FDR < 0.05), we identified the score with the greater incremental improvement over the baseline model. The proportion of scores with greater incremental improvement among AFR and EUR populations is plotted. Error bars represent 95% confidence intervals for the proportion.

